# Mortality in hemodialysis: Synchrony of biomarker variability indicates a critical transition

**DOI:** 10.1101/2021.05.14.21257235

**Authors:** Alan A. Cohen, Diana L. Leung, Véronique Legault, Dominique Gravel, F. Guillaume Blanchet, Anne-Marie Côté, Tamàs Fülöp, Sylvia Juhong Lee, Frédérik Dufour, Mingxin Liu, Yuichi Nakazato

## Abstract

Critical transition theory suggests that complex systems should experience increased temporal variability just before abrupt change, such as increases in clinical biomarker variability before mortality. We tested this in the context of hemodialysis using 11 clinical biomarkers measured every two weeks in 763 patients over 2496 patient-years. We show that variability – measured by coefficients of variation – is more strongly predictive of mortality than biomarker levels. Further, variability is highly synchronized across all biomarkers, even those from unrelated systems: the first axis of a principal component analysis explains 49% of the variance. This axis then generates powerful predictions of all-cause mortality (HR95=9.7, p<0.0001, where HR95 is a scale-invariant metric of hazard ratio across the predictor range; AUC up to 0.82) and starts to increase markedly ∼3 months prior to death. Such an indicator could provide an early warning sign of physiological collapse and serve to either trigger intervention or initiate discussions around palliative care.

## Main

In many medical contexts, apparently stable patients decompensate or decline relatively rapidly. Examples include epileptic seizures^1^, decompensation in intensive care^2^, and clinical frailty in the elderly, which represents a point of accelerated decline^3,4^. These events represent critical transitions – abrupt shifts of a complex system from one attractor state to another^5,6^. Work from complex systems theory on ecosystems, economies, and climate regimes has shown that critical transitions have very similar properties in apparently distinct systems^7,8^, and that they can be predicted by using appropriate metrics of their dynamics known as early warning signs (EWSs) characterizing increases in variance^9,10^, change in auto-correlation structure^3,11^, and increases in cross-correlation^5^, all of which are part of a phenomenon known as “critical slowing down” that precedes the transition^12^. EWSs of critical transitions could allow for corrective action, particularly to avoid system collapse^13^. Several studies have started to successfully apply critical transition theory in a medical context^3,14–16^, but have not yet fully drawn on the multivariate nature of biological systems. Multivariate indices of EWSs might successfully incorporate not just the dynamics of individual biomarkers preceding transitions, but also the joint dynamics of the numerous interacting biomarkers, substantially increasing predictive power.

We propose that beyond specific diseases^17–19^, the broad health status of an individual can be assessed using critical transition theory, with general outcomes such as death often reflecting a critical transition, which we refer to as a “physiological collapse” of the organism. Successful prediction of impending physiological collapse could allow timely interventions to reduce mortality and/or start end-of-life planning. However, one challenge in a medical context is obtaining sufficiently detailed time-series data to calculate meaningful EWSs. Clinical data are often subject to reporting biases (e.g., measures taken when patients are sick, selective reporting of diagnostic codes), and cohort studies are rarely if ever at a fine enough temporal scale. An exception is hemodialysis data in patients with end-stage renal disease: in most contexts, hemodialysis occurs 3x/week with blood draws approximately bi-weekly, continuing over several years, until kidney transplant or death^20^. Accordingly, data on hemodialysis patients present a time series of largely complete biomarker data generally unavailable in other medical contexts, and permit us to assess changes in variability in ways that are generally not possible in cohort data or other standard clinical contexts. Recently, albumin variability was shown to increase before death in a Japanese cohort of hemodialysis patients^20^, and a multivariate index of variability predicted frailty in the same cohort^21^. Here, using mortality as a proxy for physiological collapse in hemodialysis patients, we predicted that the variability of biomarkers would provide a stronger prediction of impending mortality than the levels of the biomarkers. We likewise predicted that the general behavior of the system, as characterized by joint (multivariate) signals of the markers, would give stronger predictions than the levels or variability of any individual marker. Accordingly, we expect integrative measures of variability to be powerful EWSs. We assessed (1) whether there is a generalized joint signal of biomarker variability (i.e., synchrony of variability), and (2) if any such signal could predict critical transitions, in our case study, death.

## Results

### Study population and biomarker selection

We studied biomarker trajectories in all patients with chronic kidney disease on long-term dialysis in the Eastern Townships region of Quebec, based on data extracted from a clinical research database containing electronic patient records (see online Methods for details). After exclusions, our study population consisted of 763 patients (Table 1, Fig. 1) followed for a total of 56,019 visits with biomarker data over 2496 patient-years (median [interquartile range]: 45 visits/patient [15, 101] and 2.1 years/patient [0.7, 4.4]). 525 patients died during follow-up, but 91 of these had no visit within 3 months prior to their death, likely due to transfer to palliative care. Patients excluded from analyses (Fig. 1) were less likely to have diabetes, die, or have a kidney transplant, and were followed for a shorter time period (Supplementary Table 1). We detected 391/763 (51.2%) of patients as diabetic. We generated two lists of biomarkers based on their measurement frequency: one that includes 11 biomarkers generally measured every two weeks (2-week list), and one including 16 biomarkers measured every four months (4-month list, see Table 1). Main analyses were conducted with the 2-week list, while we used the 4-month list to validate our results at a different time scale and incorporate albumin, which was only measured every four months in our cohort.

**Table 1.** Study population characteristics at first visit included in analyses.

| Characteristic | (n = 763) | 2-week list | 4-month list |
| --- | --- | --- | --- |
| Age (years), mean ± SD | 64.2 ± 15.8 | | |
| Age range (years), min - max | 16.2 - 94.6 |  |  |
| Men, n (%) | 479 (62.8) |  |  |
| Diabetics, n (%) | 391 (51.2) |  |  |
| Death, n (%) | 525 (68.8) |  |  |
| Kidney transplant (KT) |  |  |  |
| Patients who ceased HD after having a KT, n (%)<sup>a</sup> | 93 (12.2) |  |  |
| Total number of patients with KT, n (%) | 186 (24.4) |  |  |
| Follow-up length (years), median (Q25, Q75) | 2.1 (0.7, 4.4) |  |  |
| Albumin (g/L), mean ± SD | 36.1 ± 5.9 | | X |
| Calcium (mmol/L), mean ± SD | 2.25 ± 0.19 | | X |
| Creatinine (μmol/L), mean ± SD | 667 ± 303 | | X |
| Glucose (mmol/L), mean ± SD<sup>b</sup> | 8.1 ± 5.0 | | X |
| Hematocrit (proportion of 1), mean ± SD | 0.32 ± 0.05 | X | X |
| Hemoglobin (g/L), mean ± SD | 108 ± 15 | X | X |
| MCH (pg), mean ± SD | 31.3 ± 2.2 | X | X |
| MCHC (g/L), mean ± SD | 333 ± 11 | X | X |
| MCV (fL), mean ± SD | 94.0 ± 6.1 | X | X |
| Phosphate (mmol/L), mean ± SD | 1.67 ± 0.53 | | X |
| Platelet count ($10^9$/L), mean ± SD<sup>c</sup> | 215 ± 80 | X | X |
| Potassium (mmol/L), mean ± SD | 4.78 ± 0.74 | X | X |
| RBC count ($10^{12}$/L), mean ± SD | 3.5 ± 0.5 | X | X |
| RDW (%), mean ± SD<sup>b</sup> | 15.5 ± 1.9 | X | X |
| Sodium (mmol/L), mean ± SD | 138 ± 4 | X | X |
| WBC count ($10^9$/L), mean ± SD<sup>b</sup> | 7.8 ± 3.3 | X | X |

<sup>a</sup> Within 30 days from the kidney transplant.
<sup>b</sup> Variable was log-transformed.
<sup>c</sup> Variable was square root-transformed.
Abbreviations: KT, kidney transplant; MCH, mean corpuscular hemoglobin; MCHC, mean corpuscular hemoglobin concentration; MCV, mean corpuscular volume; Q25, 25<sup>th</sup> percentile; Q75, 75<sup>th</sup> percentile; RBC, red blood cells; RDW, red cell distribution width; SD, standard deviation; WBC, white blood cell.

**Figure 1.**
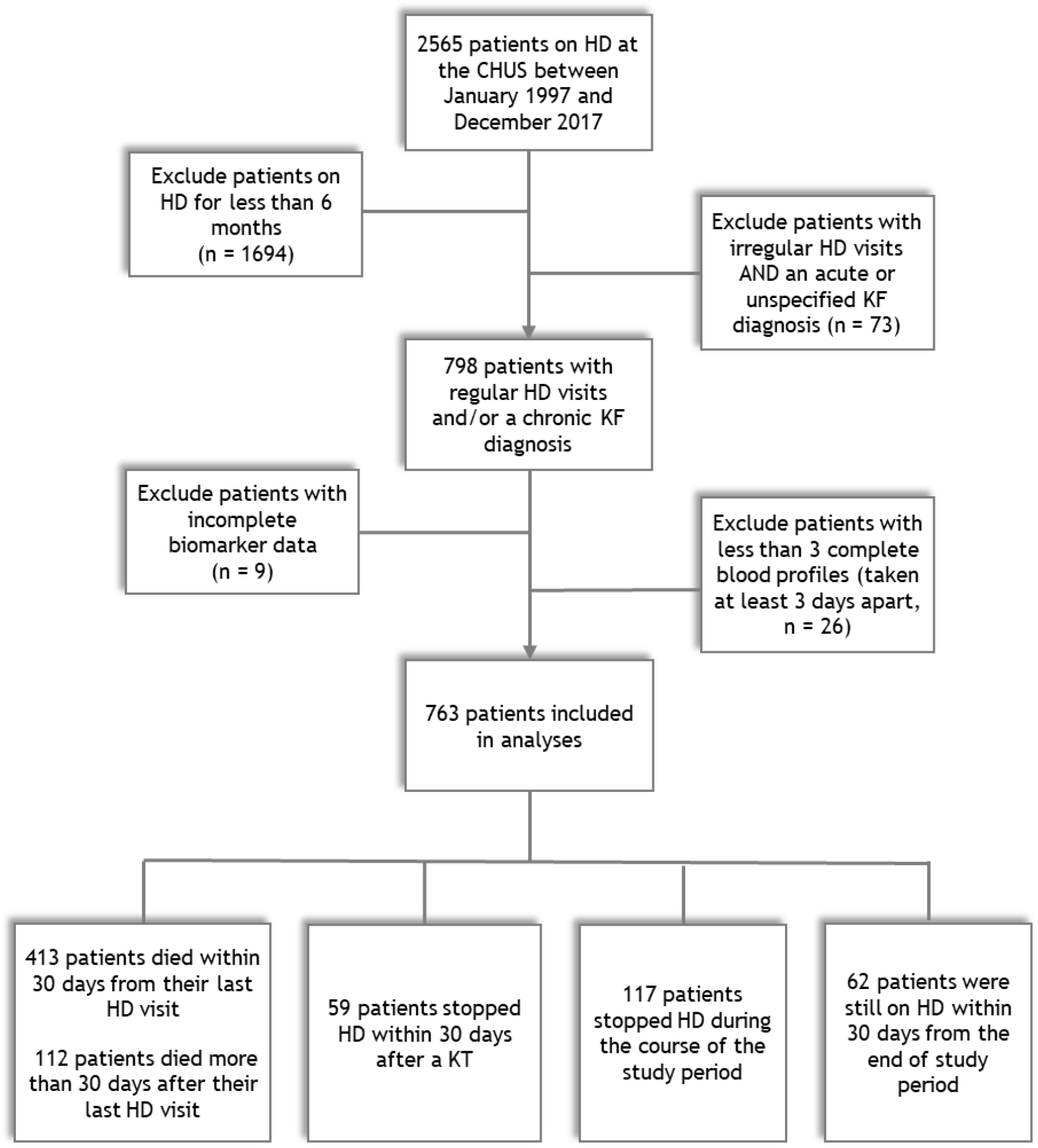
Study population flowchart diagram. Of the 2565 patients having hemodialysis (HD) at the CHUS between January 1997 and December 2017, we excluded 1694 patients who had HD for less than six months and 73 patients who had both an irregular HD visit frequency AND an acute or unspecified kidney failure (KF) diagnosis. Then, we selected visits with biomarker data at least three days apart, and excluded 9 patients with incomplete biomarker data (including 4 without any biomarker profiles) and 26 patients with less than three visits in total. The final study population thus included 763 patients, among whom 413 died within 30 days from their last HD visit and 112 who died more than 30 days after their last HD visit. Within patients still alive at the end of the study period, 59 had a kidney transplant (KT) after which they stopped HD (within 30 days), 117 stopped HD for unknown reasons, and 62 were still on HD within 30 days from the end of the study period.

### Prediction of mortality by levels and variability of individual biomarkers

First, we tested the hypothesis that individual biomarker variability would better predict mortality than individual biomarker levels. Variability of individual biomarkers was calculated as within-individual coefficients of variation (CVs) per year before death or censoring. To ensure comparability (since CVs, unlike biomarker levels, cannot be calculated at a specific timepoint), levels of biomarkers were calculated as within-individual means per year. We thus hereafter refer to information on levels as “means,” just as variability is referred to as “CVs.” We compared indices using Cox proportional hazards models to extract HR95, a scale-invariant hazard ratio for continuous variables that assesses variation in hazard across the range of the variable^22^. Overall, CVs were better in predicting mortality compared to means: all 11 biomarkers were predictive of mortality when using their CVs and nine had HR95 ≥ 4 (Fig. 2A). In comparison, nine biomarkers predicted mortality significantly when using their levels, but only one had HR95 ≥ 4 (red cell distribution width [RDW], HR95 = 4.96, *p* < 0.001; see Fig. 2A). For 10 of 11 biomarkers, the HR95 estimated from CVs was greater than that estimated from levels (Fig. 2A).

**Figure 2.**
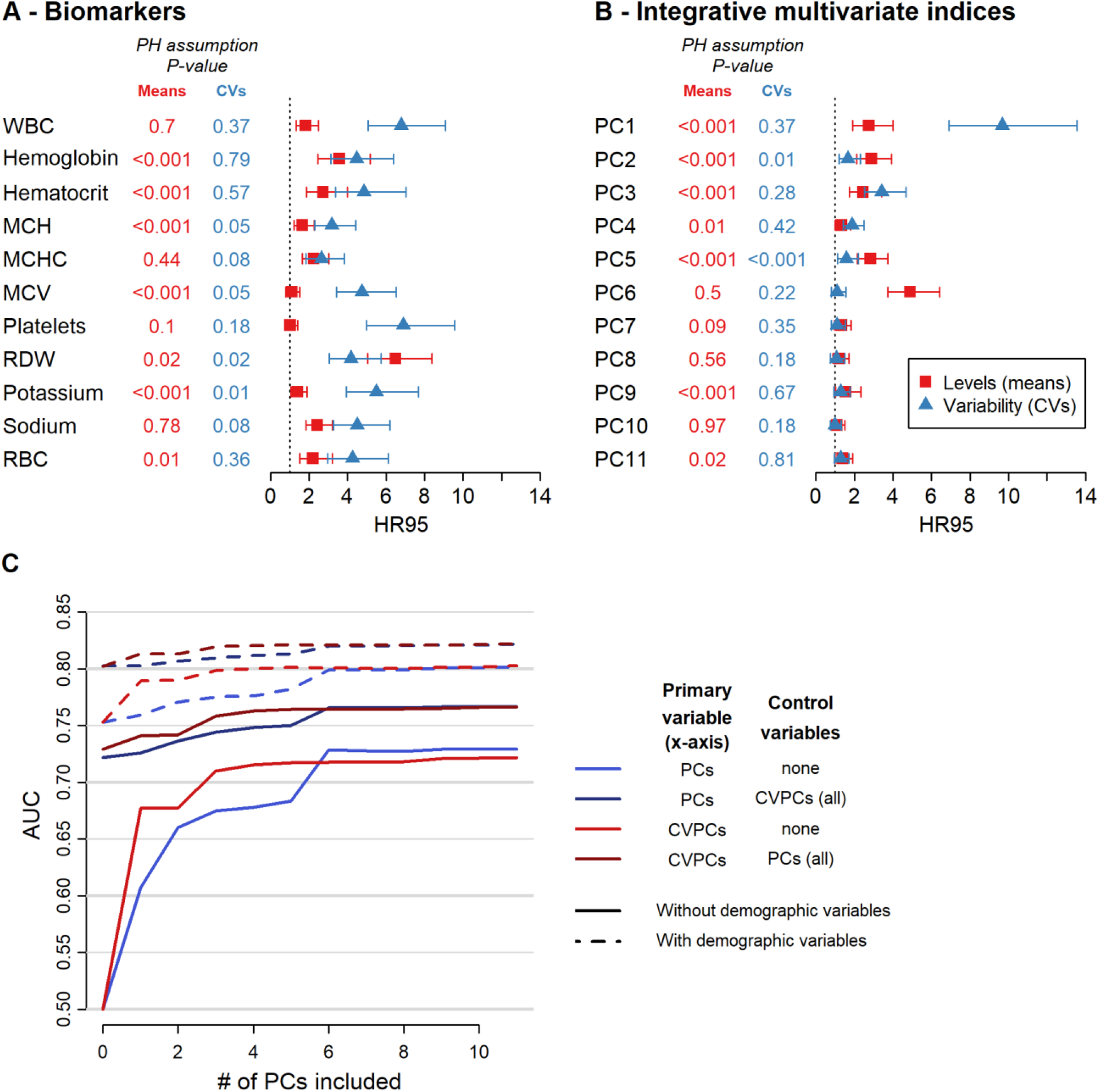
Prediction of death by biomarker levels and variability indices. **A-B**, HR95, i.e. the hazard ratio of being in the 97.5^th^ percentile relative to the 2.5^th^ percentile of the index, together with 95% confidence intervals are shown for the levels (means, red) and variability (CVs, blue) of each biomarker considered (**A**) and integrative multivariate indices (i.e. each principal component calculated on all biomarkers, **B**). All models control for age (using a cubic spline), sex, diabetes diagnosis, and length of follow-up, clustering multiple observations per individual. Levels of hemoglobin, hematocrit, MCH, MCHC, MCV, potassium, sodium, and RBC were inversed (1/x) to obtain HR95 above 1, for ease of representation. P-values of proportional assumption tests for the given coefficients are indicated. **C**, Accuracy of mortality prediction for the first principal component of a PCA performed on means (PC1, blue) or CVs (CVPC1, red), or on either one controlling for the other PCs/CVPCs in the cox model (darker hues), by sequentially increasing the number of PCs/CVPCs added in the cox model. Cox proportional hazard models were performed with (dashed lines) or without (solid lines) including demographic control variables, namely age (modelled as a cubic spline), sex, diabetes diagnosis, and length of follow-up. P-values of proportional assumption tests for the given coefficients are indicated. See Supplementary Fig. 5 for results with the 4-month variable list. Abbreviations: AUC, area under the curve; MCH, mean corpuscular hemoglobin; MCHC, mean corpuscular hemoglobin concentration; MCV, mean corpuscular volume; RBC, red blood cells; RDW, red cell distribution width; WBC, white blood cells.

### Integration of individual biomarkers into multivariate indices

To test the hypothesis that integrative measures would outperform univariate ones, we then combined information from individual biomarkers into integrative indices using principal component analysis (PCA), both for biomarker means and CVs, and extracted principal components (hereinafter referred to as “PCs” for means and “CVPCs” for CVs). The first principal component for biomarker variability (CVPC1) was by far the strongest predictor of mortality (HR95 = 9.68, *p* < 0.001, after controlling for age, sex, diabetes diagnosis, and length of follow-up), although several other indices also provided reasonably strong predictions (e.g., PC6 for biomarker mean levels, HR95 = 4.89; see Fig. 2B). Results were similar when excluding 112 patients who died but were not followed in the last 30 days before their death. In this case CVPC1 was even slightly more predictive (HR95 = 11.49, *p* < 0.001, see Supplementary Fig. 1). Compared to other predictive indices, CVPC1 was also the most stable across population subsets, with HR95 > 5 in all subgroups tested (Supplementary Fig. 2).

Next, we assessed whether prediction could be improved by combining information from our two types of integrative indices (means and CVs), by sequentially combining all PCs or CVPCs one by one (Fig. 2C). The area under the curve (AUC) increased from 0.677 to 0.741 when adding all 11 PCs to CVPC1 in the model, and reached 0.813 when including covariates (sex, age, diabetes diagnosis, and length of follow-up). However, adding subsequent CVPCs to the model had only a moderate effect on prediction accuracy (AUC = 0.822 compared to 0.813), with CVPC3 having the most effect (Fig. 2C and Supplementary Fig. 3). Broadly, the AUCs show that there are independent contributions of information from biomarker levels (means), variability (CVs), and demographic data, with the CVs contributing more than the means and CVPC1 providing by far most of the signal among CVPCs.

### CVPC1 as a joint signal of biomarker variability

Because CVPC1 provided such strong prediction of mortality, we took a closer look at what it quantifies. CVPC1 captured an impressive proportion of overall system variability, explaining 48.8% of total CV variance, whereas the second and third principal components (CVPC2 and CVPC3) explained only ∼12% and ∼10% of the variance, respectively (Fig. 3A). Most strikingly, CVPC1 is driven jointly by all biomarkers rather than a subset: Loadings of individual biomarker CVs were nearly equal, a result highly replicable across men, women, diabetics, non-diabetics, and groups at various time intervals before death (Fig. 3B). Supporting this, all pairwise correlations among biomarker CVs were positive, statistically significant at α = 0.05 (Fig. 3C), and generally of similar magnitude (Fig. 3D). These results show that variability is highly synchronized across the individual biomarkers: as one biomarker became more variable, all the others tend to be as well. In contrast, biomarker means were much less synchronized than CVs: PC1, which by definition explains the most variance of any axis in the analysis, explained only ∼25% of total variance in biomarker levels, and contributions of individual biomarkers were much less equally distributed (Figs. 3A, 3C, and Supplementary Fig. 4).

**Figure 3.**
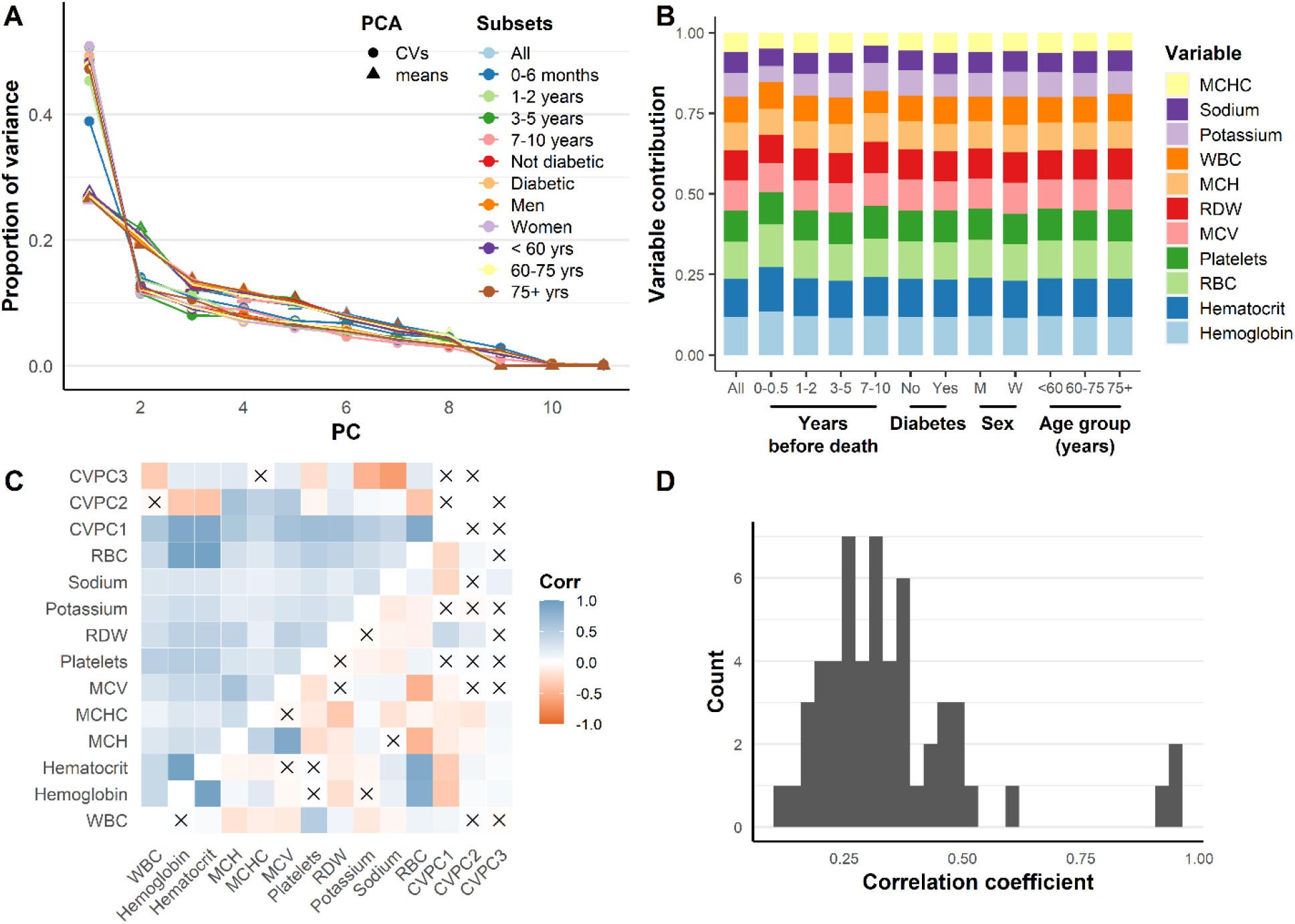
Physiological variability shows a strong coordinated signal distributed evenly across all measured biomarkers. **A**, Variance explained by PCA on raw biomarkers (triangles) or coefficients of variation (circles), for different population subsets relative to time of death or by demography. Note that variance is more concentrated in the first axis for CVs relative to raw variables. **B**, Relative biomarker contributions to CVPC1, ordered from largest contribution (hemoglobin) to smallest (MCHC) in the full dataset (see online Methods). Subsequent columns are based on loadings of the PCA run exclusively on the indicated subsets. Contribution for a given biomarker is the absolute value of the loading divided by the sum of the absolute values of all loadings. Note that contributions are nearly equal for all biomarkers and are highly stable across all population subsets. **C**, Pearson correlations (Corr) among raw biomarkers, coefficients of variation, and composite indices. CVPC1-3: First through third axes of the PC on coefficients of variation. Blue indicates positive correlations, and red represents negative correlations. Xs represent correlations not significant at α=0.05. Above the diagonal are the CVs, and below are the biomarker levels. Note that all 55 correlations among CVs are statistically significant. **D**, Histogram of correlation coefficients between CVs of individual biomarkers showing relatively little variation in the strength of correlations (mean *r*= 0.35, SD = 0.18, min = 0.12, max = 0.96). See Supplementary Fig. 6 for results with the 4-month variable list. Abbreviations: MCH, mean corpuscular hemoglobin; MCHC, mean corpuscular hemoglobin concentration; MCV, mean corpuscular volume; RBC, red blood cells; RDW, red cell distribution width; WBC, white blood cells.

### Validation using the 4-month biomarker list

Albumin, whose variability was previously found to predict mortality in hemodialyzed patients^20^, was only measured every four months in our study population. Therefore, we replicated key analyses using the 4-month list of 16 biomarkers, for which a total of 11,475 visits were available, with a median [interquartile range] of 8 [3, 18] visits per patient (see Supplementary Figs. 5-9). Results were broadly similar, though with somewhat smaller effect sizes (e.g., HR95 = 4.99, *p* < 0.001 for CVPC1, Supplementary Fig. 5), showing the stability of the patterns regardless of precise biomarker choice or temporal scale of the measurement.

### Timing of change in CVPC1 before death

Finally, we assessed how long before death the joint variability increase could be detected by applying change point analysis to our integrative measure of biomarker variability (CVPC1) calculated with different time windows (i.e., every 2, 3, or 4 months). A stable change point was detected at ∼3 months before death (Fig. 4A). This rise in CVPC1 before death, which is more abrupt than for any of the other PCs or CVPCs (Supplementary Fig. 10), is also weakly discernible in randomly chosen individual profiles, and even prior to hospitalization events (Supplementary Fig. 11). Biomarker levels increase or decrease at different rates preceding death (Fig. 4B), whereas increases in CVs appear to be synchronized in the few months preceding death (Fig. 4C). Albumin, which is only in the 4-month list, appears to show the greatest increase in variability in the year preceding death (Supplementary Fig. 12), while among biomarker levels, RDW shows the greatest increase before death in both the 2-week and the 4-month list (Fig. 4B).

**Figure 4.**
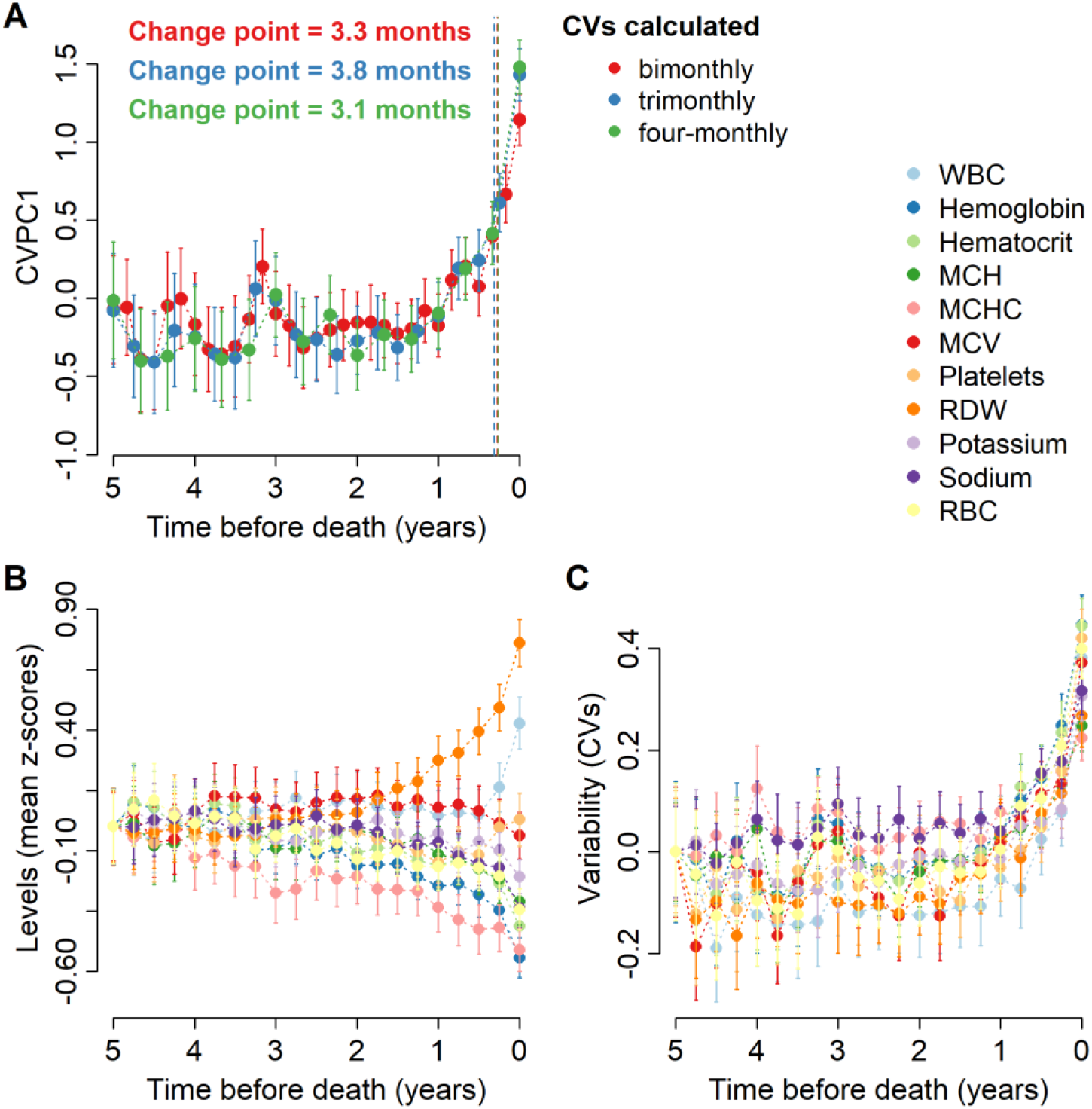
CVPC1 trend before death. **A**, CVPC1, the first principal component of a PCA performed on CVs calculated every 2 (red), 3 (blue), or 4 (green) months, is plotted against time before death. A change point analysis – applied to regression models between CVPC1 and time before death and allowing slopes to vary across individuals – revealed a change in CVPC1 distributions at ∼ 3 months before death, represented by the vertical dashed lines with the respective colors. **B-C**, Biomarker levels (mean z-scores) and variability (CVs) were calculated every 3 months and averages are plotted against time before death. For ease of comparison, means and CVs were centered at 5 years before death. See Supplementary Fig. 12 for results with the 4-month list. Abbreviations: MCH, mean corpuscular hemoglobin; MCHC, mean corpuscular hemoglobin concentration; MCV, mean corpuscular volume; RBC, red blood cells; RDW, red cell distribution width; WBC, white blood cells.

## Discussion

In this study we have shown that a sudden, coordinated rise in the variability of biomarkers is a strong predictor of impending mortality. Mortality is notoriously challenging for clinicians to predict more than a few days in advance; our index can detect 11-fold differences in mortality risk, with indications visible ∼3 months before death. More broadly, the variability of biomarkers is more predictive than their levels, and integrative indices are more predictive than individual biomarker variation. Interestingly, the single biomarker showing the highest predictive performance and pre-mortem increase is RDW, a finding consistent with previous studies^23–25^. RDW is itself a kind of CV – quantified through a simple equation (i.e. [standard deviation of red blood cell volumes] / [mean corpuscular volume] × 100) – and can thus be regarded as an indicator of momentary variability.

These findings also have broad theoretical implications. They confirm that critical transition theory can apply to health, not only in highly specific clinical contexts, but also to overall health/homeostasis and its converse, physiological collapse. We also report a striking synchronization in the increase in variability across biomarkers: even biomarkers that do not correlate with each other show synchronized increases in variability (Figs 3 and 4C). In other words, the signal of the impending critical transition propagates quickly and diffusely across the whole system. While this propagation is consistent with complex systems theory, its biological underpinnings are not yet known. Nevertheless, such propagation across unrelated biomarkers is consistent with recent findings suggesting that loss in resilience (frailty) emerges from parallel dysregulation in different physiological systems^4,26,27^. Indeed, complex systems theory states that system resilience decreases before a critical transition, whereby the decrease in resilience is characterized by slower rates in recovery after disturbances, and thus higher variability^28^.

The timescales of years used in this study are long relative to many other physiological critical transitions. It might be expected, for example, that similar signals could be observed over hours or days in intensive care units with different indices^2^. Long timescales also mean we cannot distinguish whether there are truly joint shifts in biomarker variability, or whether there are ordered cascades of effects at shorter timescales. Nonetheless, for the expected rates of change in hemodialysis patients, the increase 3 months prior to death suggests a nearly ideal window for either clinical interventions to prevent decline, or for end-of-life discussions.

It should be possible to construct global indices of variability that could serve as useful clinical predictors, particularly in combination with other data (mean values, demographics, etc.). Here, our best models achieved an AUC of 0.82, a reasonable prediction by most standards. In this context, an exceptional number (e.g., 0.95) would be nearly impossible to obtain since all - cause mortality is very broad and should not be precisely predictable on timescales of years. We are using mortality as an admittedly imperfect proxy for physiological collapse, and we hypothesize that our indices would predict a direct measure of physiological collapse substantially better than they predict all-cause mortality.

While this study shows the clinical potential of biomarker variability as an EWS, more work is required before clinical implementation. Other multivariate indices of variability exist beyond CVPC1^29^, and variability is only one signal among many that can be extracted from time series (e.g. lag-1 autocorrelations, flickering, skewness, etc.^28^). Also, here we could only include albumin in the 4-month list due to its measurement frequency in our study population, but our results suggest that if it were measured every two weeks it might substantially improve the performance of integrative measures. Future work will need to compare and likely integrate multiple approaches, validating optimal prediction algorithms before clinical testing; our emphasis here is on demonstrating the potential of such methods.

It is not yet clear whether clinical interventions could successfully rescue patients from physiological collapse, though apparent spikes in CVPC1 before non-fatal hospitalizations (Supplementary Fig. 11) suggest hope in at least some cases. Alternatively, the best clinical application of EWSs may be in triggering discussions on end-of-life care preferences with patients. Access to quality palliative care and medical assistance in dying is becoming a priority^30,31^, and identifying the right time and way to initiate such a discussion still appears to be challenging for many nephrologists^32,33^. Once a sufficient algorithm and its application are validated, it should be possible to integrate into electronic health data systems^34^.

Here we have shown applications of biomarker dynamics to hemodialysis, but many other medical applications could readily be developed, including vital rates in intensive care^2^, brain waves to predict sleep cycles^35^, and advent of clinical frailty^3^. Several machine learning-based algorithms exist^36–38^, including to predict mortality in chronic kidney disease patients^39–41^; however, our model is one of the few to rely on clear clinical constructs. With more regular data collection, there could also be applications in congestive heart failure, predicting myocardial infarction, chronic obstructive pulmonary disease, and many others, as well as to fields such as ecology, economics, and climate science. Current prediction of critical transitions, both in medical practice and across disciplines, is largely based on level or dynamics of one or two biomarkers that attempt to summarize the state of a complex system. Integration across multiple biomarkers is likely to better capture underlying processes^42^, as we have shown here.

In conclusion, our findings have important implications at three levels. At a theoretical level, we showed surprising synchronicity in the variability of all measured biomarkers, implying that compartmentalization of physiology breaks down drastically preceding adverse critical transitions. At a methodological level, this suggests that the principles we illustrated can be leveraged to develop similarly powerful predictive indices in many other medical contexts – intensive care, diabetes, heart failure, and cancer, to name a few. At a practical clinical level, our findings imply that powerful predictors of mortality risk in hemodialysis patients can be extracted from electronic medical record systems easily and within an appropriate time frame to help clinicians consider interventions or initiate a timely discussion with patients on end-of-life care.

## Methods

### Study design and setting

This is a retrospective cohort study to aiming to apply critical transition theory in a clinical context – hemodialysis – to test whether increased biomarker variability predicts mortality. The study took place at the CHUS hospital (*Centre hospitalier universitaire de Sherbrooke*) in Sherbrooke (Quebec, Canada).

### Ethical approval and patient consent

This project received ethical approval by the *Comité d’éthique de la recherche du CIUSSS de l’Estrie – CHUS* (#2015-788, 14-059). Written informed consent for participation was not required for this study in accordance with the National Legislation and the Institutional Requirements, due to the retrospective nature of the study.

### Participants and data sources

Our study population consisted of all chronic kidney disease (CKD) hemodialysis patients at the CHUS hospital in Sherbrooke, Quebec, Canada between 1997 and 2017. CHUS protocol is hemodialysis 3x/week with a fixed blood work panel every two weeks to ensure follow-up and adjust treatment^20^. Patient records were extracted from the CIRESSS (*Centre informatisé de recherche évaluative en services et soins de santé*) database, which aggregates all electronic hospital data for clinical and administrative purposes since 1991, a nearly exhaustive sample for the Eastern Townships region of Québec, population ∼326,000. To capture patients on whom sufficient data were gathered and with CKD rather than acute kidney failure, from 2565 initial patients, we excluded 1694 patients who were no longer treated by in-center hemodialysis after six months (indicating death, recovery within six months from acute kidney failure or transition to another renal replacement modality or to conservative care), and 73 patients with irregular hemodialysis visits and/or an acute or unspecified kidney failure diagnosis. This left 798 patients with CKD on long-term hemodialysis. We excluded 28,681 visits (30%) with missing data (Fig. 1 and Supplementary Table 2). Because the start of hemodialysis itself may represent a critical transition^43^, we also excluded data for the first six months on hemodialysis for each patient; however, for 26 patients, we were left with less than three visits and thus excluded these patients. Our final study cohort was comprised of 763 patients, for whom we included all visits for which biomarker data were gathered between earliest available hemodialysis visit (after the initial six - month exclusion) and death or last hemodialysis visit (Table 1).

### Biomarkers and other key variables

We included all biomarkers measured regularly in the context of hemodialysis (Table 1), and excluded those measured only irregularly or having an important proportion of missingness in our cohort (uric acid, urea, iron binding capacity, iron, ferritin, iron saturation, transferrin, carbon dioxide, partial carbon dioxide pressure, partial oxygen pressure, pH, ionized calcium, intact parathyroid hormone, glycated hemoglobin and thyroid-stimulating hormone). For truncated laboratory variables (i.e. fields containing “<” or “>” signs), we generated the remaining part of the distribution (the tails of the distribution) using the `rtnorm` function from the `MCMCglmm` package (version 2.29)^44^. Mortality was identified via hospital and provincial (*Régie d’assurances maladie du Québec*) death records. Hospitalisation dates were recorded. Diabetes status was identified based on diagnostic codes 250 for ICD 9 before April 2006, and E10, E11, E13, E14 for ICD-10 after April 2006.

### Missing data

To assess if data in our study cohort were missing completely at random, we performed Kruskal - Wallis test for age and Pearson’s Chi-squared test for discrete covariates (sex, diabetes, and mortality), comparing observations with complete vs incomplete biomarker data. The significant differences in covariates suggest that our data are not missing completely at random (Supplementary Table 2). Nevertheless, imputation was considered, but not used, for the following reasons:

1. Imputation methods have not been validated in this context, in contrast to more standard applications such as linear regression.
2. A successful imputation would have to preserve not only expected mean value, but expected variance of the ensemble, across relevant time periods. While it might be possible to develop methods that could achieve this, it would be a major undertaking in itself.
3. The creation of a potentially valid imputation method would require us to make numerous assumptions that might or might not be respected; as such, the risk of inducing a bias in the imputation is substantially larger than the risk of a bias due to the missing data.
4. If the result with imputed data conflicted with the current result, we would have no way of knowing for certain which method was preferable, but for reason 3) we would revert to the original analysis anyway.
5. The validation of the method and the stability of the results across numerous, distinct population subsets gives a strong indication that the result is not a function of population composition, which is the primary concern that would motivate imputation in this case.

### Calculation of Early Warning Signs

Biomarkers were log-transformed for normality as appropriate (Table 1). To test differences between biomarker levels and variability, and between univariate versus multivariate indices, we constructed time series for four types of EWSs calculated by individual across time periods counting backwards from death/censoring: 1) levels of individual biomarkers were calculated as within-individual means per time period; 2) variability of individual biomarkers was calculated as within-individual CVs per time period; 3) multivariate (integrative) indices of biomarker levels (PCs) were generated using PCA applied on within-individual biomarker means; and 4) multivariate (integrative) indices of biomarker variability (CVPCs) were generated using PCA applied on within-individual CVs. Unless otherwise specified (e.g., for change-point analysis with bimonthly, trimonthly, or four-monthly calculation), PCA were conducted on CVs or means calculated by year working backwards from time of death or censoring. Analyses were replicated in subpopulations to assess generalisability of findings: not diabetics (n=372), diabetics (n=391), men (n=479), women (n=284), <60 years old (n=241), 60 to 75 years old (n=368), and 75+ years old (n=280). Age was defined based on age at visit, so some individuals were found in different categories over time.

To calculate PCs based on biomarker means, we ran a PCA on all centered and scaled biomarker raw values using `prcomp` function in R, extracted the loadings, and applied them to the within-individual biomarker means calculated by year, thereby generating scores for each PC for every patient across years. CVs were calculated for each biomarker in each individual at each time period and were log-transformed. In a few cases, especially for tightly regulated biomarkers such as sodium and potassium, the CV per year was equal to zero because of laboratory rounding; for ease of analysis, we replaced these zeros by 90% of the next smallest CV value. We ran a PCA on the adjusted CVs (see below) using `prcomp` function in R, centering and scaling them.

### Adjustment of CVs for the number of observations included in the calculation

We noticed that CVs were biased by the number of values included in their calculation (Supplementary Fig. 13A). To confirm that this is a property of CV calculation, rather than of our data, we explored this association in simulated data. Simulated data were generated for each biomarker (transformed if needed to meet the assumption of normality) with the `rnorm` function, using the mean and standard deviation from our study population. Here, we only present results from one biomarker, but they all behave the same way: the fewer simulated values included in CV calculation, the lower CVs tended to be (Supplementary Fig. 13B). To control for this bias in CV calculation, we used the residuals from nonlinear regression models (CV=((1)/(√(n))×a+b), performed separately for each variable with the `nls` function in R).

These residuals were used in the PCA to calculate CVPCs. We also ran sensitivity analyses using only CVs calculated with at least five observations, and using them directly in the PCA, instead of correcting with the non-linear regression (Supplementary Fig. 13C-D). These sensitivity analyses show that our results are strongly robust to these specific analytic details.

### Mortality prediction

Power of various EWSs to predict death was assessed using counting process Cox proportional hazards models^45,46^. We used the hazard ratios to calculate “HR95” as a way to compare effect sizes across variables of varying scales: it represents the hazard ratio for an individual in the 97.5^th^ percentile of the variable relative to one in the 2.5^th^, and permits apples-to-apples comparison across alternative biomarker indices on different scales. We assessed mortality risk with the `coxph` function (`survival` package version 3.1-12)^47^, clustering multiple observations per individual using the “cluster” argument. Demographic covariates include age (modeled as a cubic spline with five degrees of freedom (df) using the `bs` function from the `splines` package to better account for non-linear relationships with age), sex, diabetes diagnosis, and years of follow-up. All models included the square root of observation number included in CVPC calculation as weights to account for the lower precision in CV estimation with fewer observations included in its calculation (note that the non-linear regression mentioned above does not correct for the loss of precision; it corrects for the bias in CV calculation leading to smaller CV values when fewer observations are available, not for the precision). We calculated the AUC with the `pROC` package (version 1.16.2) using a non-parametric method^48^, while the Cox proportional hazards assumption was verified using the `cox.zph` function.

#### Change point analysis

We assessed the timing of changes in our integrative measures of biomarker variability (CVPCs) prior to death using change point regression with the `mcp` package (version 0.3.0)^49^. Regression models were performed on all available CVPC values, and slopes were allowed to vary across individuals, with results presented after averaging values by time point.

## Data Availability

Raw data presented in this article cannot be made publicly available, due to confidentiality concerns, transfer, and sharing of individual-level data. Data used in this study require prior approval from the CIRESSS and the Director of Professional Services of the CHUS, as well as by the Comite d'Ethique de la Recherche du CIUSSS de l'Estrie - CHUS. Requests to access the datasets should be directed to https://www.crchus.ca/en/services-outils/autresservices-et-outils/infocentre/.

## Acknowledgements

This work was supported by the Canadian Institutes of Health Research (CIHR, grant #153011 awarded to AAC) and by the Fonds de recherche du Québec – Société et culture (FRQSC, grant 2020-AUDC-270433 awarded to AAC, DG, FGB, and TF). AAC is supported by a Fonds de recherche du Québec – Santé (FRQ-S) Senior Salary Award and is a member of the FRQ-S funded *Centre de recherche du CHUS* and *Centre de recherche sur le vieillissement*.

## Contributions

A.A.C., D.L.L., V.L., D.G., G.B., and T.F. conceived the study and designed the analyses. S.J.L., D.L.L., and V.L. contributed to data acquisition and preparation. D.L.L. and V.L. performed statistical analyses, while F.D. independently replicated main findings. A.A.C., T.F., A.-M.C., and Y.N. contributed to interpretation of clinical aspects of results. A.A.C. and V.L. wrote the first draft of the manuscript and all authors critically revised the manuscript.

## Data availability

Raw data presented in this article cannot be made publicly available, due to confidentiality concerns, transfer, and sharing of individual-level data. Data used in this study require prior approval from the CIRESSS and the Director of Professional Services of the CHUS, as well as by the *Comité d’Éthique de la Recherche du CIUSSS de l’Estrie* – CHUS. Requests to access the datasets should be directed to https://www.crchus.ca/en/services-outils/autresservices-et-outils/infocentre/.

## Code availability

All analyses were performed using the R statistical language (versions 3.6.0 and 4.0.0)^50^ and main findings were replicated independently by one co-author (F.D.) to assure replicability. Code is available at github.com/cohenaginglab/HD_variability.

## References

1. Kramer, M. A. et al. Human seizures self-terminate across spatial scales via a critical transition. Proc. Natl. Acad. Sci. U. S. A. (2012) doi:10.1073/pnas.1210047110.

2. Ghalati, P. F. et al. Critical Transitions in Intensive Care Units: A Sepsis Case Study. 1902.05764, (2019).

3. Gijzel, S. M. W. et al. Dynamical Resilience Indicators in Time Series of Self-Rated Health Correspond to Frailty Levels in Older Adults. Journals Gerontol. - Ser. A Biol. Sci. Med. Sci. 72, 991–996 (2017).

4. Fried, L. P. et al. The physical frailty syndrome as a transition from homeostatic symphony to cacophony. Nat. Aging 1, 36–46 (2021).

5. Scheffer, M. et al. Early-warning signals for critical transitions. 461, 53–59 (2009).

6. Scheffer, M. Complex systems: Foreseeing tipping points. Nature (2010) doi:10.1038/467411a.

7. May, R. M., Levin, S. A. & Sugihara, G. Complex systems: Ecology for bankers. Nature (2008) doi:10.1038/451893a.

8. Lenton, T. M., Livina, V. N., Dakos, V., Van Nes, E. H. & Scheffer, M. Early warning of climate tipping points from critical slowing down: Comparing methods to improve robustness. Philos. Trans. R. Soc. A Math. Phys. Eng. Sci. (2012) doi:10.1098/rsta.2011.0304.

9. Carpenter, S. R. & Brock, W. A. Rising variance: A leading indicator of ecological transition. Ecol. Lett. (2006) doi:10.1111/j.1461-0248.2005.00877.x.

10. Drake, J. M. & Griffen, B. D. Early warning signals of extinction in deteriorating environments. Nature (2010) doi:10.1038/nature09389.

11. Ghanavati, G., Hines, P. D. H., Lakoba, T. I. & Cotilla-Sanchez, E. Understanding early indicators of critical transitions in power systems from autocorrelation functions. IEEE Trans. Circuits Syst. I Regul. Pap. (2014) doi:10.1109/TCSI.2014.2332246.

12. Dakos, V. & Bascompte, J. Critical slowing down as early warning for the onset of collapse in mutualistic communities. Proc. Natl. Acad. Sci. (2014) doi:10.1073/pnas.1406326111.

13. Macrae, C. Early warnings, weak signals and learning from healthcare disasters. BMJ Quality and Safety (2014) doi:10.1136/bmjqs-2013-002685.

14. Almeida, V. G. & Nabney, I. T. Early warnings of heart rate deterioration. in Proceedings of the Annual International Conference of the IEEE Engineering in Medicine and Biology Society, EMBS (2016). doi:10.1109/EMBC.2016.7590856.

15. Chen, L., Liu, R., Liu, Z. P., Li, M. & Aihara, K. Detecting early-warning signals for sudden deterioration of complex diseases by dynamical network biomarkers. Sci. Rep. (2012) doi:10.1038/srep00342.

16. Trefois, C., Antony, P. M. A., Goncalves, J., Skupin, A. & Balling, R. Critical transitions in chronic disease: Transferring concepts from ecology to systems medicine. Current Opinion in Biotechnology (2015) doi:10.1016/j.copbio.2014.11.020.

17. Tarazona, A., Forment, J. & Elena, S. F. Identifying early warning signals for the sudden transition from mild to severe tobacco etch disease by dynamical network biomarkers. Viruses (2019) doi:10.3390/v12010016.

18. Rockne, R. C. et al. State-Transition Analysis of Time-Sequential Gene Expression Identifies Critical Points That Predict Development of Acute Myeloid Leukemia. Cancer research (2020) doi:10.1158/0008-5472.CAN-20-0354.

19. Li, M., Zeng, T., Liu, R. & Chen, L. Detecting tissue-specific early warning signals for complex diseases based on dynamical network biomarkers: Study of type 2 diabetes by cross-tissue analysis. Brief. Bioinform. (2014) doi:10.1093/bib/bbt027.

20. Nakazato, Y., Kurane, R., Hirose, S., Watanabe, A. & Shimoyama, H. Aging and death - associated changes in serum albumin variability over the course of chronic hemodialysis treatment. PLoS One 12, e0185216 (2017).

21. Nakazato, Y. et al. Estimation of homeostatic dysregulation and frailty using biomarker variability: a principal component analysis of hemodialysis patients. Sci. Rep. 10, 1–12 (2020).

22. Milot, E. et al. Trajectories of physiological dysregulation predicts mortality and health outcomes in a consistent manner across three populations. Mech Ageing Dev 141–142, 56–63 (2014).

23. Mucsi, I., Ujszaszi, A., Czira, M. E., Novak, M. & Molnar, M. Z. Red cell distribution width is associated with mortality in kidney transplant recipients. Int. Urol. Nephrol. 46, 641–651 (2014).

24. Solak, Y. et al. Red cell distribution width is independently related to endothelial dysfunction in patients with chronic kidney disease. Am. J. Med. Sci. 347, 118–124 (2014).

25. Oh, H. J. et al. Red blood cell distribution width is an independent predictor of mortality in acute kidney injury patients treated with continuous renal replacement therapy. Nephrol. Dial. Transplant. 27, 589–594 (2012).

26. Ghachem, A. et al. Evidence from two cohorts for the frailty syndrome as an emergent state of parallel dysregulation in multiple physiological systems. Biogerontology 22, 63– 79 (2021).

27. Fried, L. P. et al. Nonlinear multisystem physiological dysregulation associated with frailty in older women: implications for etiology and treatment. J Gerontol A Biol Sci Med Sci 64, 1049–1057 (2009).

28. Scheffer, M. et al. Anticipating Critical Transitions. 338, (2012).

29. Liu, M. et al. Prediction of mortality in hemodialysis patients using moving multivariate distance. Front. Physiol. 12, 612494 (2021).

30. Government of Canada. Framework on Palliative Care in Canada. (2018).

31. Government of Canada. Medical assistance in dying - Canada.ca. https://www.canada.ca/en/health-canada/services/medical-assistance-dying.html#shr-pg0 (2020).

32. Borreani, C. & Miccinesi, G. End of life care preferences. Curr. Opin. Support. Palliat. Care 2, 54–59 (2008).

33. Mandel, E. I., Bernacki, R. E. & Block, S. D. Serious illness conversations in ESRD. Clin. J. Am. Soc. Nephrol. 12, 854–863 (2017).

34. Major, V. J. & Aphinyanaphongs, Y. Development, implementation, and prospective validation of a model to predict 60-day end-of-life in hospitalized adults upon admission at three sites. BMC Med. Inform. Decis. Mak. 20, 214 (2020).

35. Bashan, A., Bartsch, R. P., Kantelhardt, J. W., Havlin, S. & Ivanov, P. C. Network physiology reveals relations between network topology and physiological function. Nat. Commun. (2012) doi:10.1038/ncomms1705.

36. Horne, B. D. et al. Exceptional Mortality Prediction by Risk Scores from Common Laboratory Tests. Am. J. Med. 122, 550–558 (2009).

37. Manz, C. R. et al. Validation of a Machine Learning Algorithm to Predict 180-Day Mortality for Outpatients with Cancer. JAMA Oncol. 6, 1723–1730 (2020).

38. Wang, L. et al. Development and Validation of a Deep Learning Algorithm for Mortality Prediction in Selecting Patients with Dementia for Earlier Palliative Care Interventions. JAMA Netw. Open 2, 196972 (2019).

39. Forzley, B. et al. External validation and clinical utility of a prediction model for 6-month mortality in patients undergoing hemodialysis for end-stage kidney disease. Palliat. Med. 32, 395–403 (2018).

40. Siga, M. M. et al. Prediction of all-cause mortality in haemodialysis patients using a Bayesian network. Nephrol. Dial. Transplant. 35, 1420–1425 (2020).

41. Noh, J. et al. Prediction of the Mortality Risk in Peritoneal Dialysis Patients using Machine Learning Models: A Nation-wide Prospective Cohort in Korea. Sci. Rep. 10, 1– 11 (2020).

42. Cohen, A. A. Complex systems dynamics in aging: new evidence, continuing questions. Biogerontology 17, 205–220 (2016).

43. Broers, N. J. H., Cuijpers, A. C. M., Van Der Sande, F. M., Leunissen, K. M. L. & Kooman, J. P. The first year on haemodialysis: A critical transition. Clinical Kidney Journal (2015) doi:10.1093/ckj/sfv021.

44. Hadfield, J. D. MCMC methods for multi-response generalized linear mixed models: The MCMCglmm R package. J. Stat. Softw. 33, 1–22 (2010).

45. Andersen, P. K. & Gill, R. D. Cox’s Regression Model for Counting Processes: A Large Sample Study. Ann. Stat. 10, 1100–1120 (1982).

46. Therneau, T. M. & Grambsch, P. M. Modeling Survival Data: Extending the Cox Model. (Springer New York, 2000). doi:10.1007/978-1-4757-3294-8.

47. Therneau, T. M. Survival analysis (R package survival version 3.1-12). https://github.com/therneau/survival (2020).

48. Robin, X. et al. pROC: An open-source package for R and S+ to analyze and compare ROC curves. BMC Bioinformatics 12, 77 (2011).

49. Lindeløv, J. K. mcp: An R Package for Regression With Multiple Change Points. OSF Prepr. (2020) doi:10.31219/osf.io/fzqxv.

50. The Comprehensive R Archive Network. http://cran.r-project.org/.

