## Supplementary Information for "Mortality in hemodialysis: Synchrony of biomarker variability indicates a critical transition"

**Supplementary Table 1.** Characteristics of patients excluded from analyses at first available visit, compared to those included.

**Supplementary Table 2.** Demographic characteristic comparison between observations with complete and incomplete biomarker data.

**Supplementary Figure 1.** Prediction of death by biomarker mean and variance indices, excluding individuals who died but were not followed-up until death.

**Supplementary Figure 2.** Mortality prediction by PC1, PC2, PC6, CVPC1, and CVPC3 in different population subsets.

**Supplementary Figure 3.** Effect of combining biomarker means and CVs on prediction of mortality for PC1, PC2, PC6, and CVPC3.

**Supplementary Figure 4.** Relative biomarker contributions to PC1, PC2, PC6, and CVPC3 for the 2-week variable list.

**Supplementary Figure 5.** Prediction of death by biomarker mean and variance indices for the 4-month variable list.

**Supplementary Figure 6.** Physiological variability shows a strong coordinated signal distributed evenly across all 16 measured biomarkers.

**Supplementary Figure 7.** Mortality prediction by PC1, PC2, PC7, CVPC1, and CVPC3 in different population subsets for the 4-month variable list.

**Supplementary Figure 8.** Effect of combining biomarker means and CVs on prediction of mortality for PC1, PC2, PC7, and CVPC3 using the 4-month variable list.

**Supplementary Figure 9.** Relative biomarker contributions to PC1, PC2, PC7, and CVPC3 for the 4-month variable list.

**Supplementary Figure 10.** Trends before death of integrative multivariate indices.

**Supplementary Figure 11.** Individual CVPC1 trajectories before death or censoring.

**Supplementary Figure 12.** Mean and CV trends before death for the 4-month variable list.

**Supplementary Figure 13.** Effect of the number of observations included in CV calculation.

### Supplementary Table 1. Characteristics at first available visit of patients excluded from analyses, compared to those included.

| Characteristic | Patients included (n = 763) | Patients excluded (n = 1802) | P-value[a] |
|---|---|---|---|
| Age (years), mean ± SD | 64.2 ± 15.8 | 63.8 ± 15.5 | 0.52 |
| Age range (years), min - max | 16.2 – 94.6 | 10.5 – 92.1 | |
| Men, n (%) | 479 (62.8) | 1108 (61.5) | 0.57 |
| Diabetics, n (%) | 391 (51.2) | 656 (36.4) | <0.001 |
| Death, n (%) | 525 (68.8) | 822 (45.6) | <0.001 |
| Kidney transplant (KT) | | | |
| Patients who ceased HD after having a KT, n (%)[b] | 93 (12.2) | 53 (2.9) | <0.001 |
| Total number of patients with KT, n (%) | 186 (24.4) | 111 (6.2) | <0.001 |
| Follow-up length (years), median (Q25, Q75) | 2.1 (0.7, 4.4) | 0.07 (0.03, 0.16) | <0.001 |

[a] P-values are based on Student's t-test (age and follow-up length) or two-proportions z-test.

[b] Within 30 days from the kidney transplant.

### Supplementary Table 2. Demographic characteristic comparison between observations with complete and incomplete biomarker data.

| Characteristic | Complete (n = 66,888) | Incomplete (n = 28,681) | P-value[a] |
|---|---|---|---|
| Age, mean ± SD | 66.7 ± 15.3 | 64.1 ± 16.3 | <0.001 |
| Men, n (%) | 39,359 (58.8) | 16,346 (57.0) | <0.001 |
| Diabetes, n (%) | 34,990 (52.3) | 14,671 (51.2) | 0.001 |
| Death, n (%) | 45,900 (68.6) | 19,383 (67.6) | 0.002 |

[a] P-values are based on a Kruskal Wallis test for age and on a chi-squared test for discrete variables.

**Supplementary Figure 1. Prediction of death by biomarker mean and variance indices, excluding individuals who died but were not followed-up until death.**

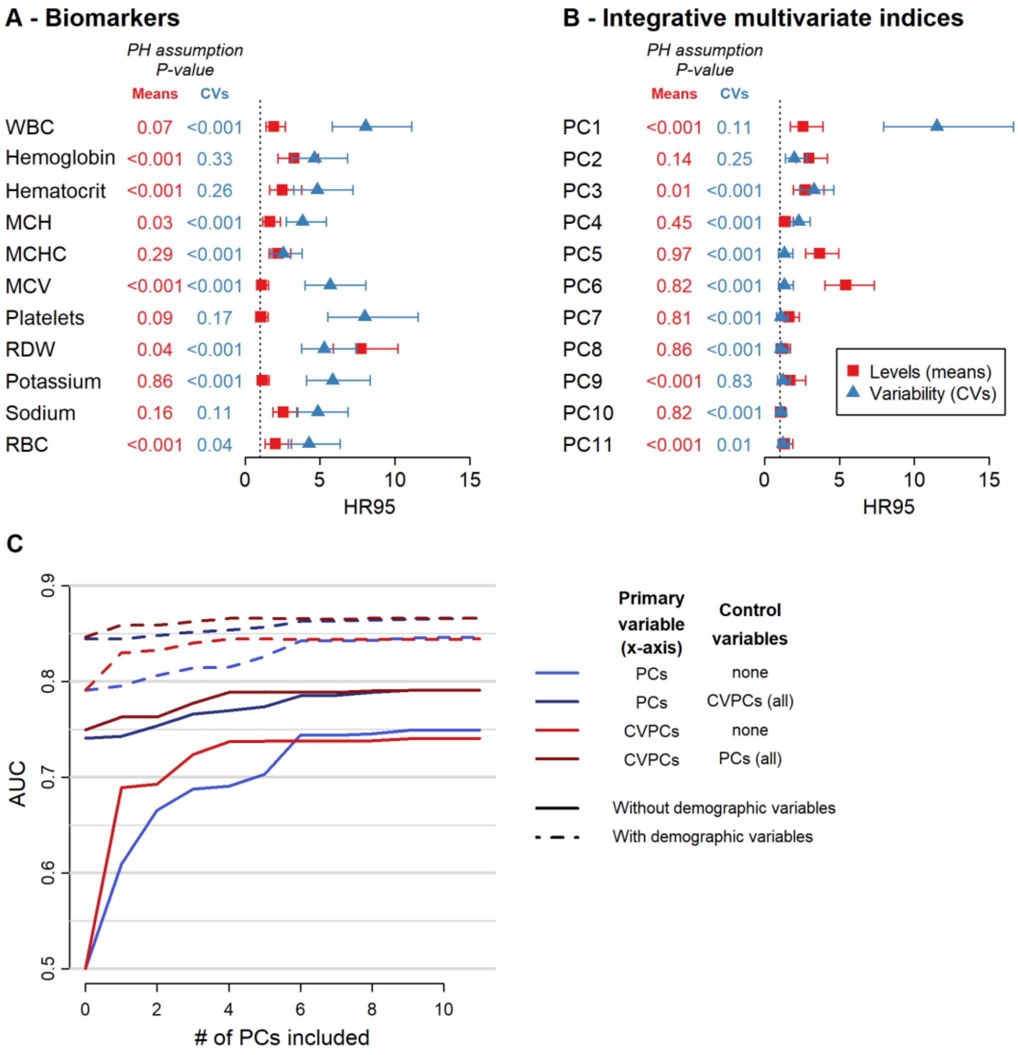


**A-B**, HR95, i.e. the hazard ratio of being in the 97.5th percentile relative to the 2.5th percentile of the index, together with 95% confidence intervals are shown for the levels (means, red) and variability (CVs, blue) of each biomarker considered (**A**), as well as for integrative multivariate indices (i.e. each principal component calculated on all biomarkers, **B**). All models control for age (using a cubic spline), sex, diabetes diagnosis, and length of follow-up, clustering multiple observations per individual, and were performed after excluding 112 individuals who died but were not followed-up until their death (n = 651). Levels of hemoglobin, hematocrit, MCH, MCHC, MCV, potassium, sodium, and RBC were inversed (1/x) to obtain HR95 above 1, for ease of representation. P-values of proportional assumption tests for the given coefficients are indicated. **C**, Accuracy of mortality prediction for the first principal component of a PCA performed on means (PC1, blue) or CVs (CVPC1, red), or on either one controlling for the other PCs/CVPCs in the cox model (darker hues), by sequentially increasing the number of PCs/CVPCs added in the cox model. Cox proportional hazard models were performed with (dashed lines) or without (solid lines) including demographic control variables, namely age (modelled as a cubic spline), sex, diabetes diagnosis, and length of follow-up. Abbreviations: AUC, area under the curve; MCH, mean corpuscular hemoglobin; MCHC, mean corpuscular hemoglobin concentration; MCV, mean corpuscular volume; RBC, red blood cells; RDW, red cell distribution width; WBC, white blood cells.

**Supplementary Figure 2. Mortality prediction by PC1, PC2, PC6, CVPC1, and CVPC3 in different population subsets.**

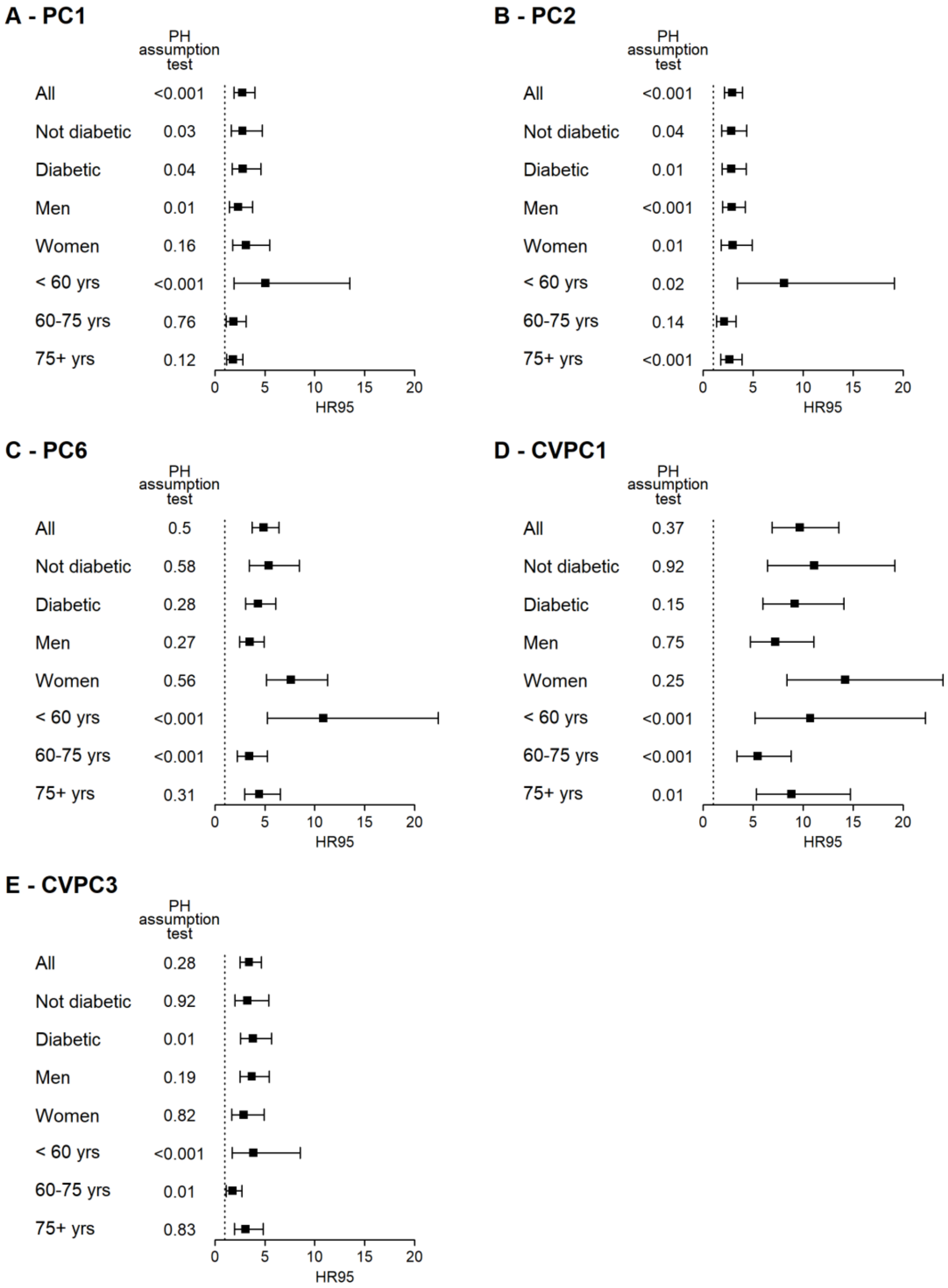


HR95, i.e. the hazard ratio of being in the 97.5th percentile relative to the 2.5th percentile of the index, together with 95% confidence intervals are shown for PC1 (**A**), PC2 (**B**), PC6 (**C**), CVPC1 (**D**), and CVPC3 (**E**) in different subsets of the study population. All models control for age using a cubic spline (with 3 degrees of freedom when performed on a specific age group and 5 otherwise) and length of follow-up, clustering multiple observations per individual. Models also control for sex and diabetes diagnosis, except when population is stratified using this variable. P-values of proportional assumption tests for the given coefficients are indicated.

**Supplementary Figure 3. Effect of combining biomarker means and CVs on prediction of mortality for PC1, PC2, PC6, and CVPC3.**

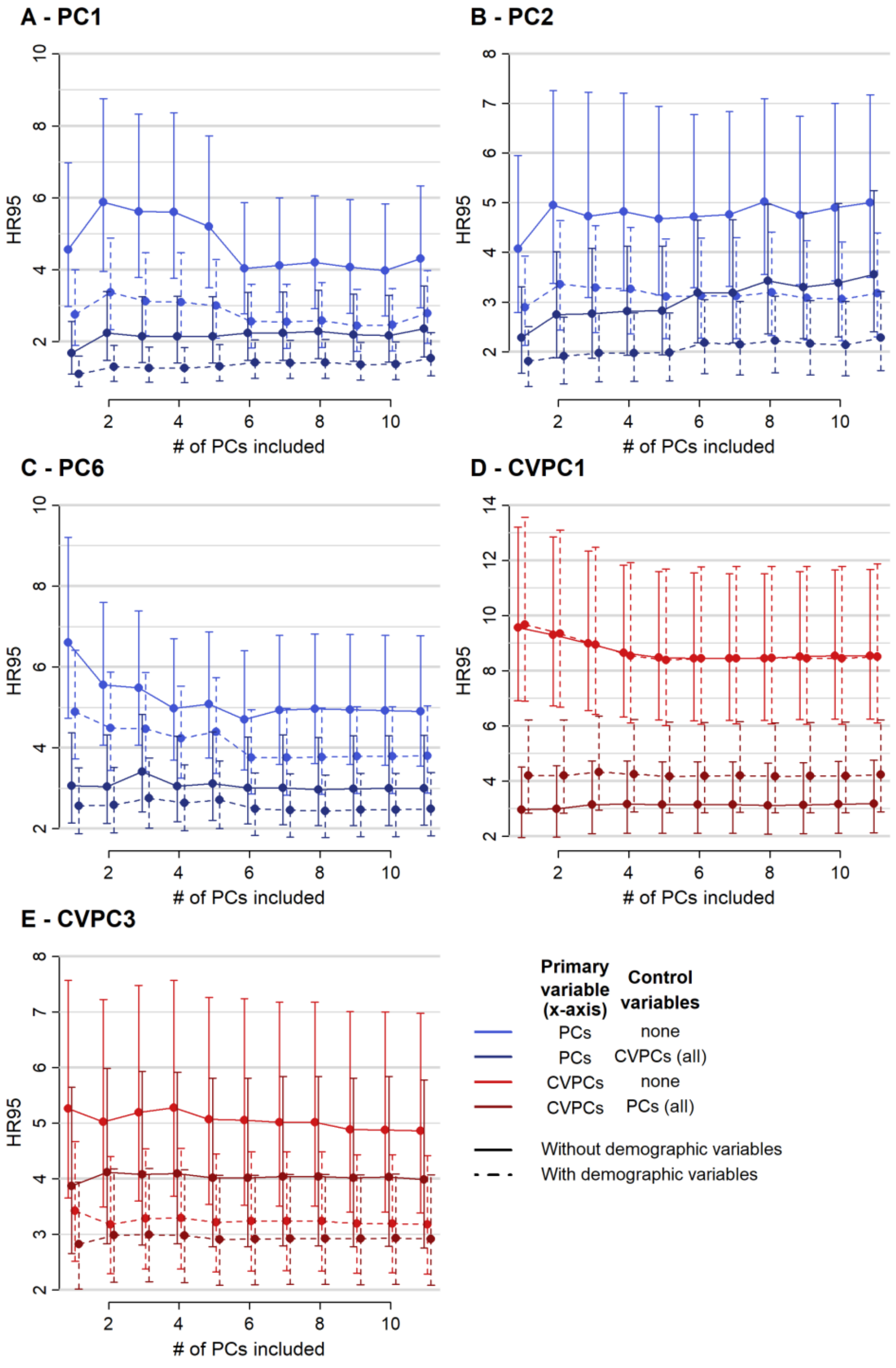


Mortality prediction for PC1 (**A**), PC2 (**B**), PC6 (**C**), CVPC1 (**D**), and CVPC3 (**E**) was assessed by sequentially adding further PCs or CVPCs to the cox model. For PC1, PC2 and PC6 (**A-C**), models including CVPCs (darker blue) and/or demographic control variables (dashed lines) were also performed. Similarly, models including PCs (darker red) and/or control variables (dashed lines) were performed for CVPC1 and CVPC3 (**D-E**). Abbreviations: CVPCs, principal components of a principal component analysis performed on biomarker coefficients of variation; HR95, difference in hazard ratio between the 97.5th and the 2.5th percentile of the index; PCs, principal components of a principal component analysis performed on raw biomarker values.

**Supplementary Figure 4. Relative biomarker contributions to PC1, PC2, PC6, and CVPC3 for the 2-week variable list.**

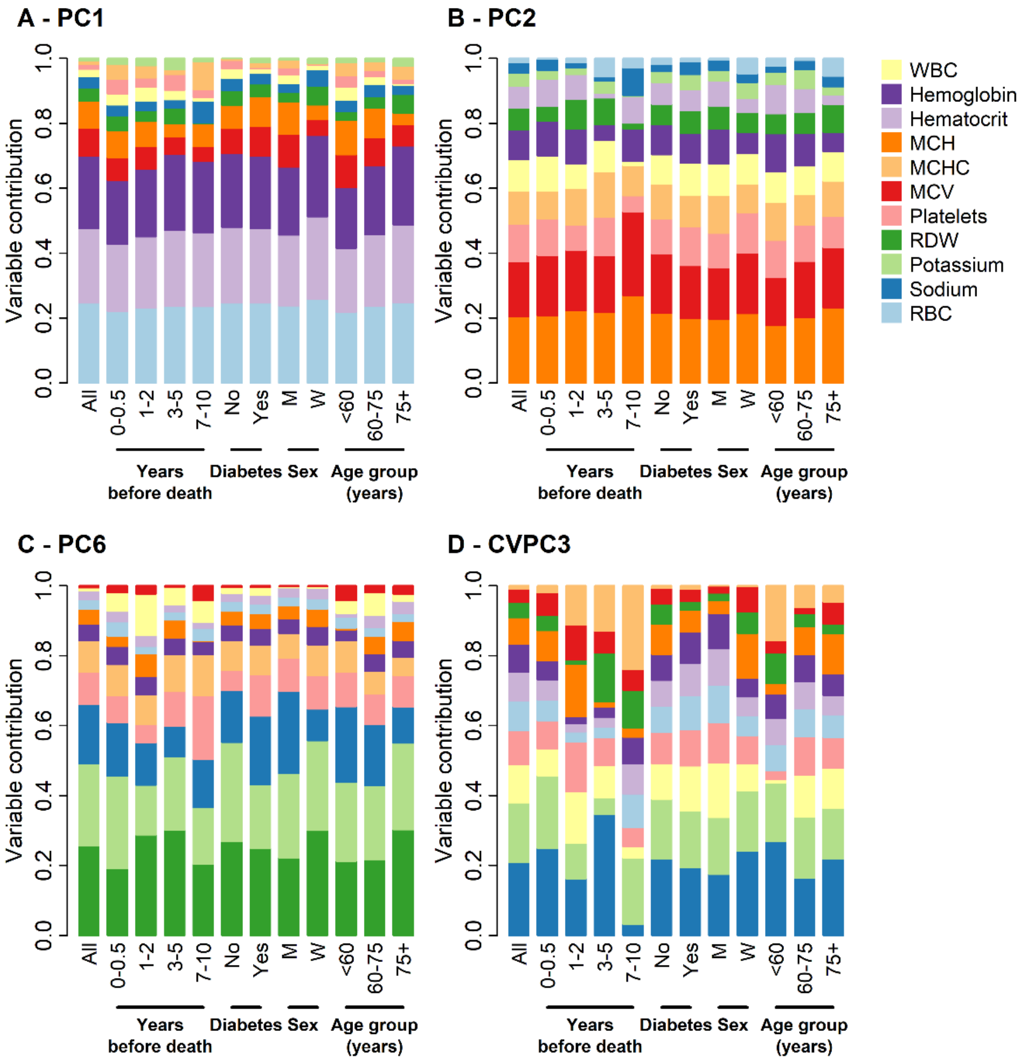


Variables are ordered from largest contribution to smallest in the full dataset; subsequent columns are based on loadings of the PCA run exclusively on the indicated subsets. Contribution for a given biomarker is the absolute value of the loading divided by the sum of the absolute values of all loadings. Variable contributions are shown for PC1 (**A**), PC2 (**B**), PC6 (**C**), and CVPC3 (**D**), which were selected based on their association with mortality risk (see Fig. 2). Abbreviations: M, men; MCH, mean corpuscular hemoglobin; MCHC, mean corpuscular hemoglobin concentration; MCV, mean corpuscular volume; RBC, red blood cells; RDW, red cell distribution width; W, women; WBC, white blood cells.

### Supplementary Figure 5. Prediction of death by biomarker mean and variance indices for the 4-month variable list.

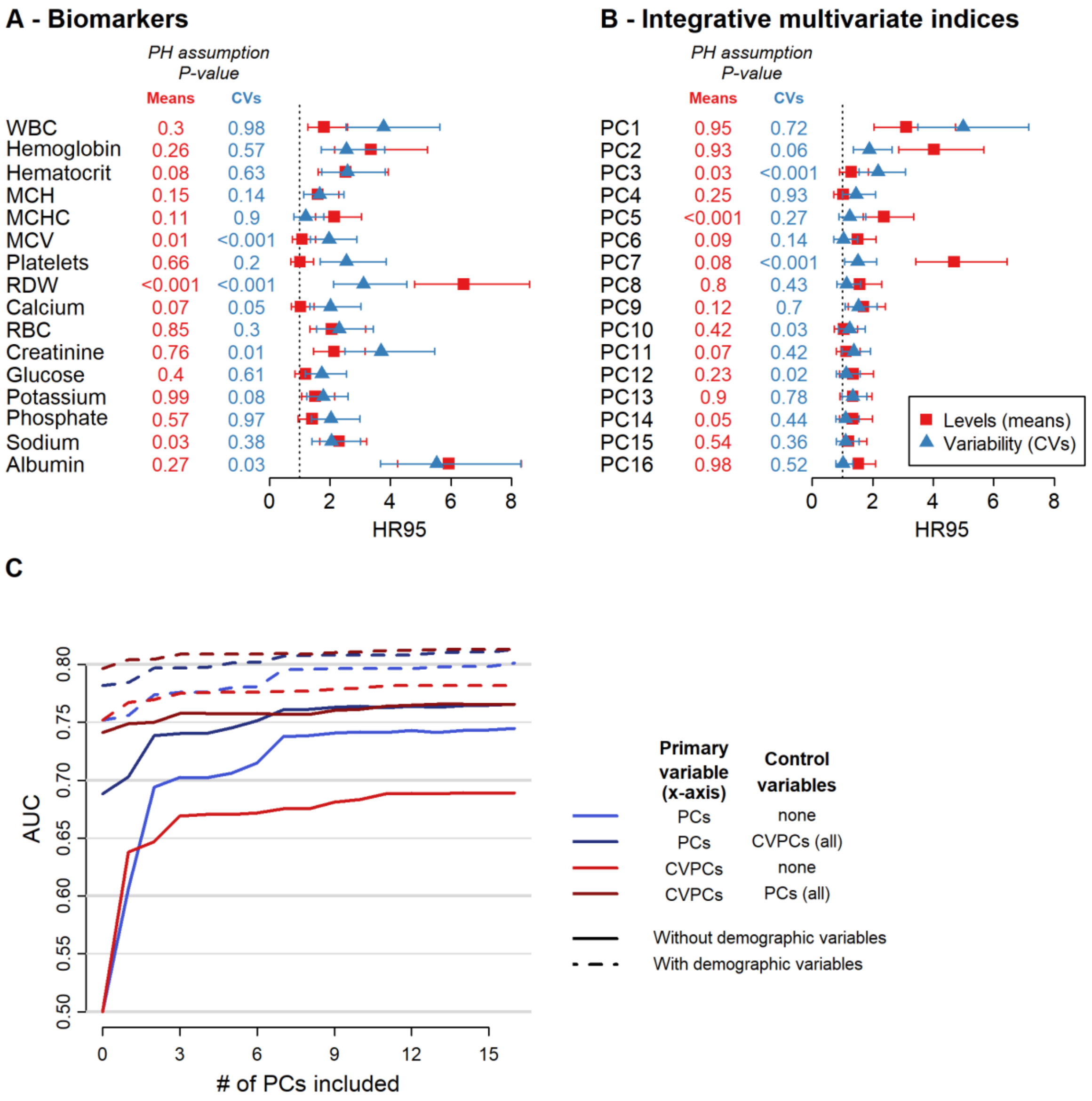


**A-B**, HR95, i.e. the hazard ratio of being in the 97.5th percentile relative to the 2.5th percentile of the index, together with 95% confidence intervals are shown for the levels (means, red) and variability (CVs, blue) of each biomarker considered (**A**), as well as for integrative multivariate indices (i.e. each principal component calculated on all biomarkers, **B**). All models control for age (using a cubic spline), sex, diabetes diagnosis, and length of follow-up, clustering multiple observations per individual. Levels of hemoglobin, hematocrit, MCH, MCHC, MCV, potassium, sodium, and RBC were inversed (1/x) to obtain HR95 above 1, for ease of representation. P-values of proportional assumption tests for the given coefficients are indicated. **C**, Accuracy of mortality prediction for the first principal component of a PCA performed on means (PC1, blue) or CVs (CVPC1, red), or on either one controlling for the other PCs/CVPCs in the cox model (darker hues), by sequentially increasing the number of PCs/CVPCs added in the cox model. Cox proportional hazard models were performed with (dashed lines) or without (solid lines) including demographic control variables, namely age (modelled as a cubic spline), sex, diabetes diagnosis, and length of follow-up. Abbreviations: AUC, area under the curve; MCH, mean corpuscular hemoglobin; MCHC, mean corpuscular hemoglobin concentration; MCV, mean corpuscular volume; RBC, red blood cells; RDW, red cell distribution width; WBC, white blood cells.

### Supplementary Figure 6. Physiological variability shows a strong coordinated signal distributed evenly across all 16 measured biomarkers.

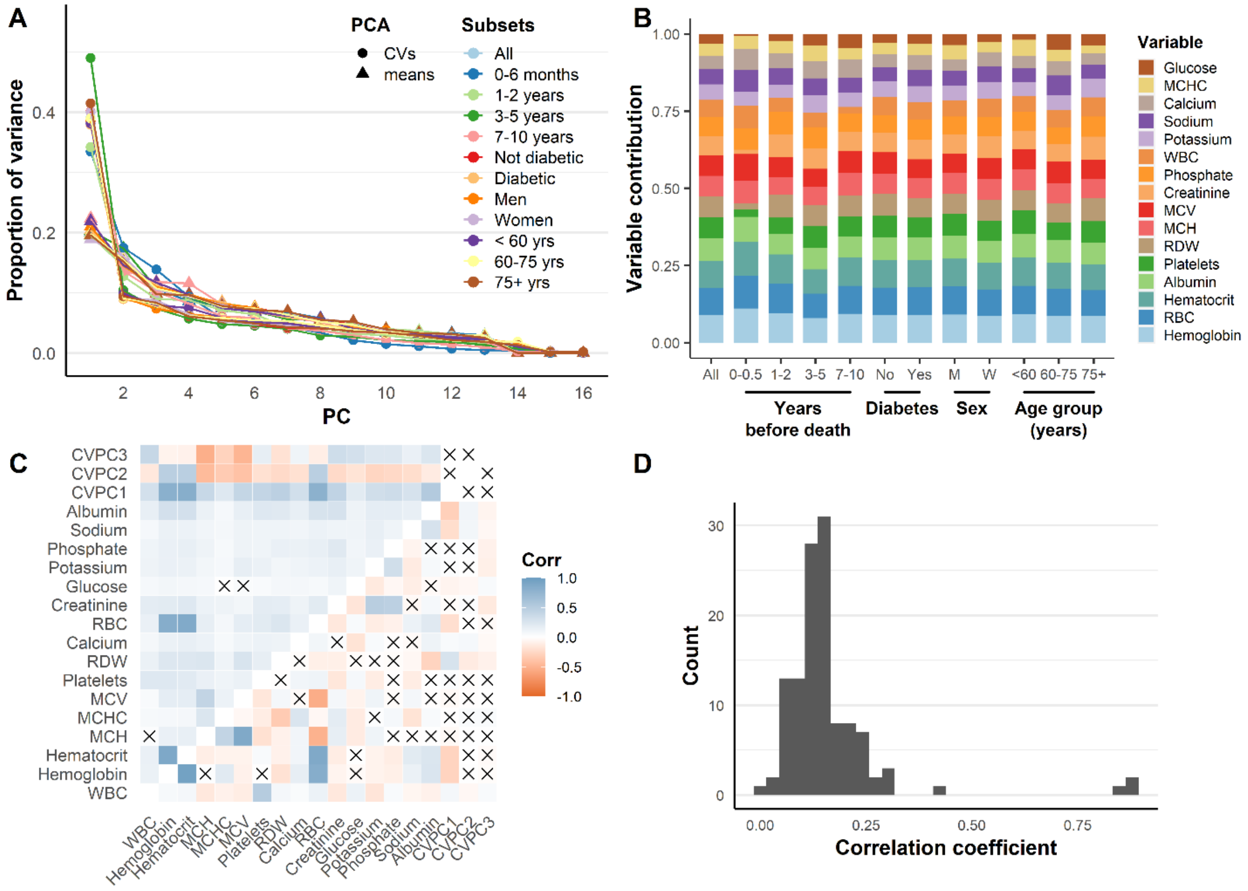


**A**, Variance explained by PCA on raw biomarkers (triangles) or coefficients of variation (circles), for different population subsets relative to time of death or by demography. Note that variance is more concentrated in the first axis for CVs relative to raw variables. **B**, Relative biomarker contributions to CVPC1, ordered from largest contribution (hemoglobin) to smallest (MCHC) in the full dataset. Subsequent columns are based on loadings of the PCA run exclusively on the indicated subsets. Contribution for a given biomarker is the absolute value of the loading divided by the sum of the absolute values of all loadings. Note that contributions are nearly equal for all biomarkers and are highly stable across all population subsets. **C**, Pearson correlations (Corr) among raw biomarkers, coefficients of variation, and composite indices. CVPC1-3: First through third axes of the PC on coefficients of variation. Blue indicates positive correlations, and red represents negative correlations. Xs represent correlations not significant at α=0.05. Above the diagonal are the CVs, and below are the biomarker levels. Note that all 120 correlations among CVs are statistically significant. **D**, Histogram of correlation coefficients between CVs of individual biomarkers showing relatively little variation in the strength of correlations (mean r= 0.16, SD = 0.13, min = 0.01, max = 0.89). Abbreviations: MCH, mean corpuscular hemoglobin; MCHC, mean corpuscular hemoglobin concentration; MCV, mean corpuscular volume; RBC, red blood cells; RDW, red cell distribution width; WBC, white blood cells.

**Supplementary Figure 7. Mortality prediction by PC1, PC2, PC7, CVPC1, and CVPC3 in different population subsets for the 4-month variable list.**

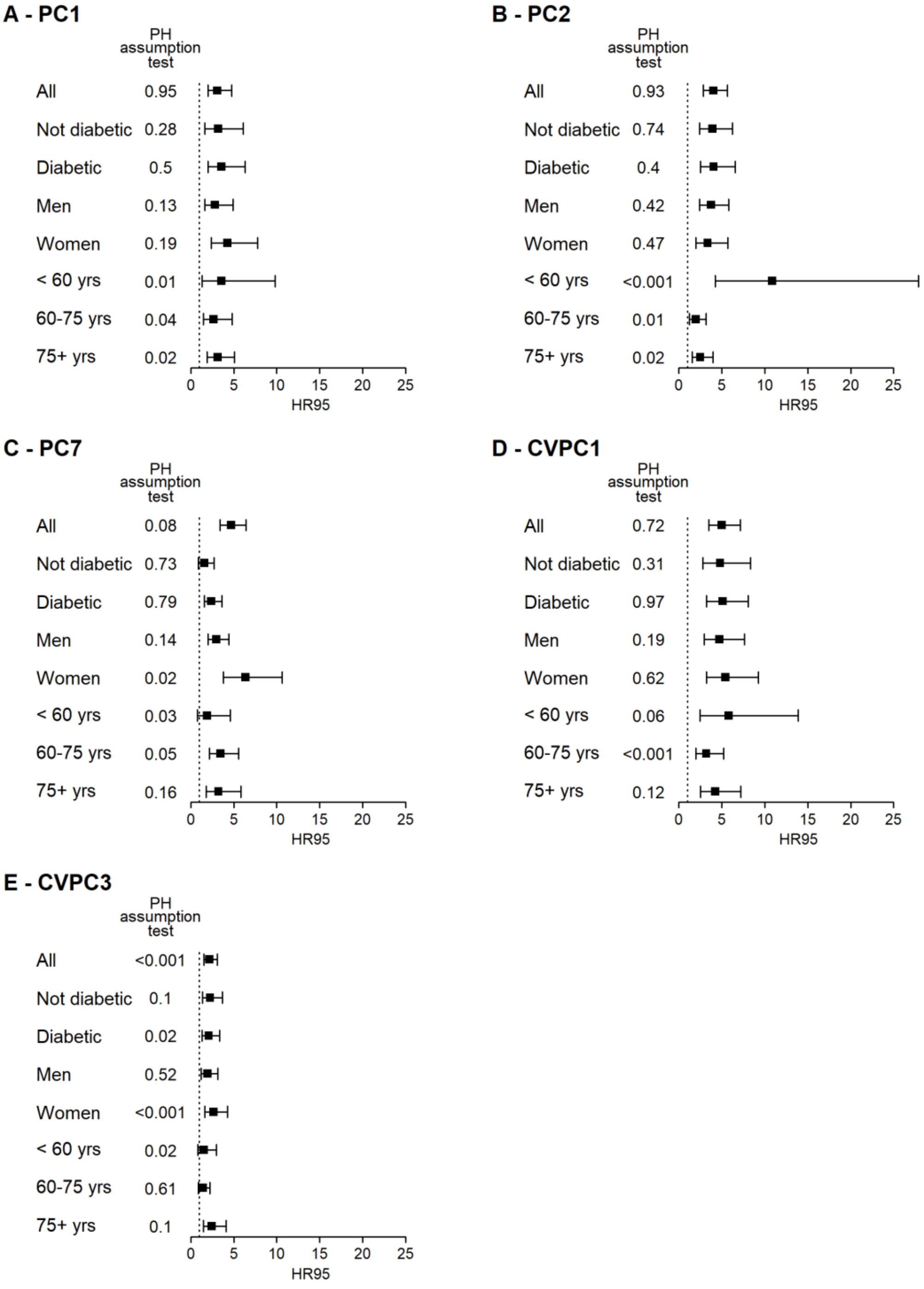


HR95, i.e. the hazard ratio of being in the 97.5$^{th}$ percentile relative to the 2.5$^{th}$ percentile of the index, together with 95% confidence intervals are shown for PC1 (**A**), PC2 (**B**), PC7 (**C**), CVPC1 (**D**), and CVPC3 (**E**) in different subsets of the study population for the 4-month variable list. All models control for age using a cubic spline (with 5 degrees of freedom when including all age groups and with 3 otherwise) and length of follow-up, clustering multiple observations per individual. Models also control for sex and diabetes diagnosis, except when population is stratified using this variable. P-values of proportional assumption tests for the given coefficients are indicated.

**Supplementary Figure 8. Effect of combining biomarker means and CVs on prediction of mortality for PC1, PC2, PC7, and CVPC3 using the 4-month variable list.**

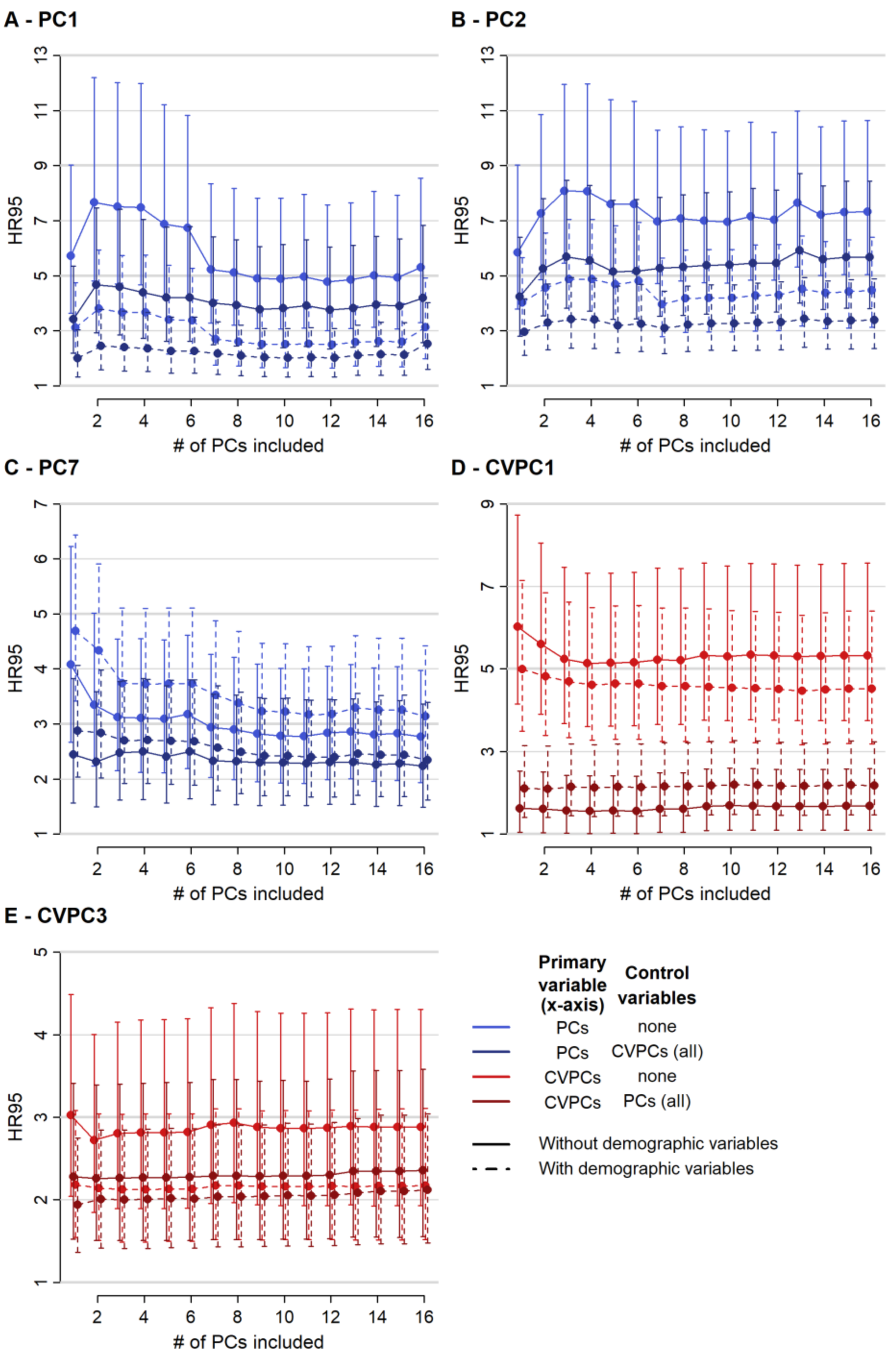


Mortality prediction for PC1 (**A**), PC2 (**B**), PC7 (**C**), CVPC1 (**D**), and CVPC3 (**E**) was assessed by sequentially adding further PCs or CVPCs to the cox model. For PC1, PC2 and PC7 (**A-C**), models including CVPCs (darker blue) and/or demographic control variables (dashed lines) were also performed. Similarly, models including PCs (darker red) and/or control variables (dashed lines) were performed for CVPC1 and CVPC3 (**D-E**). Abbreviations: CVPCs, principal components of a principal component analysis performed on biomarker coefficients of variation; HR95, difference in hazard ratio between the 97.5th and the 2.5th percentile of the index; PCs, principal components of a principal component analysis performed on raw biomarker values.

**Supplementary Figure 9. Relative biomarker contributions to PC1, PC2, PC7, and CVPC3 for the 4-month variable list.**

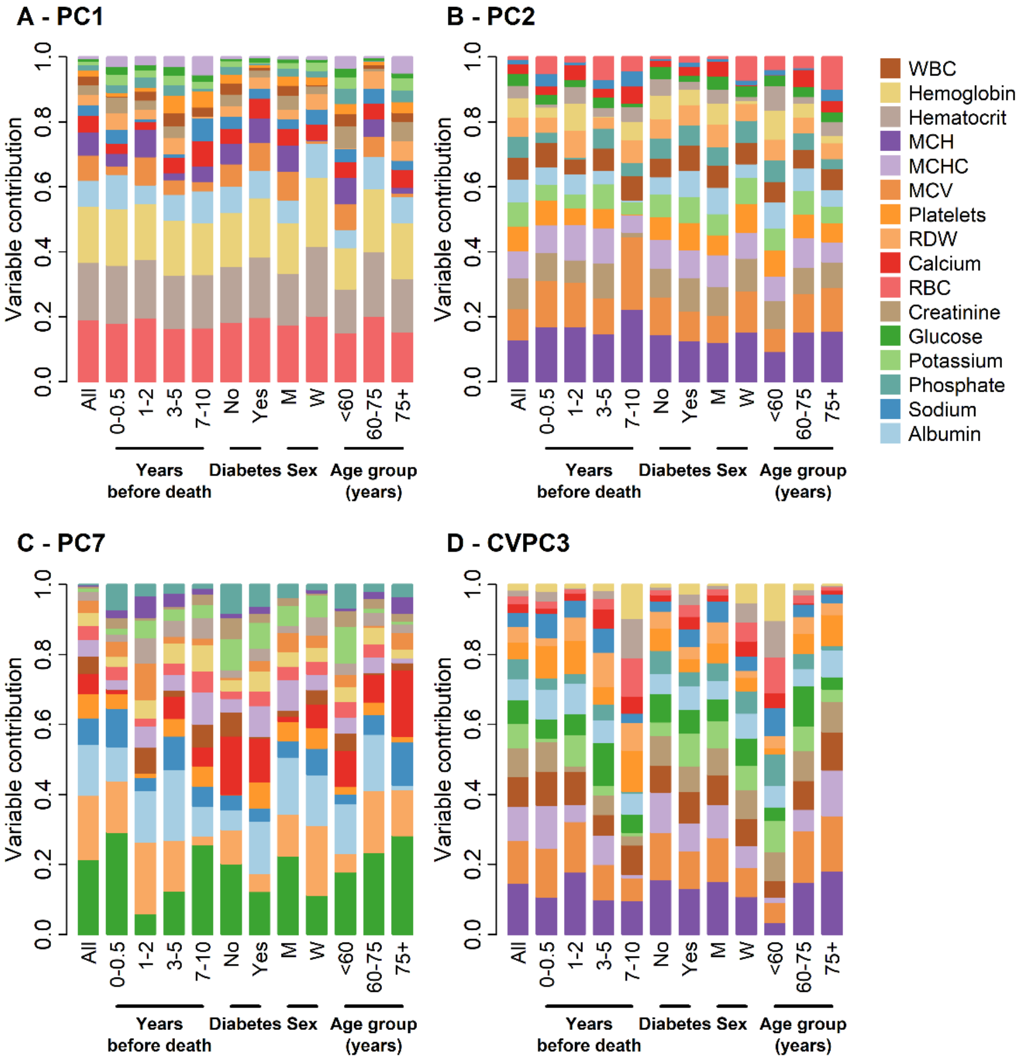


Variables are ordered from largest contribution to smallest in the full dataset; subsequent columns are based on loadings of the PCA run exclusively on the indicated subsets. Contribution for a given biomarker is the absolute value of the loading divided by the sum of the absolute values of all loadings. Variable contributions are shown for PC1 (**A**), PC2 (**B**), PC7 (**C**), and CVPC3 (**D**), which were selected based on their association with mortality risk (see Supplementary Fig. 5). Abbreviations: M, men; MCH, mean corpuscular hemoglobin; MCHC, mean corpuscular hemoglobin concentration; MCV, mean corpuscular volume; RBC, red blood cells; RDW, red cell distribution width; W, women; WBC, white blood cells.

**Supplementary Figure 10. Trends before death of integrative multivariate indices.**

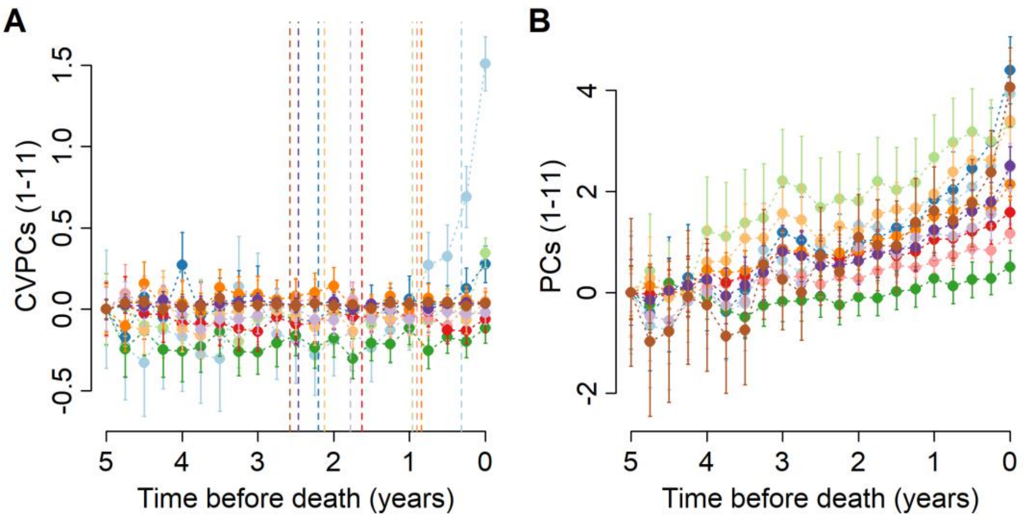


| | Change point |
|---|---|
| CVPC1 | 0.31 (0.00 - 0.73) |
| CVPC2 | 2.20 (1.02 - 2.98) |
| CVPC3 | 0.96 (0.26 - 1.69) |
| CVPC4 | 0.90 (0.23 - 1.59) |
| CVPC5 | 0.90 (0.04 - 1.52) |
| CVPC6 | 1.63 (0.67 - 2.54) |
| CVPC7 | 2.12 (1.25 - 2.72) |
| CVPC8 | 0.84 (0.00 - 1.98) |
| CVPC9 | 1.78 (0.93 - 2.87) |
| CVPC10 | 2.47 (1.73 - 3.59) |
| CVPC11 | 2.58 (1.45 - 3.46) |

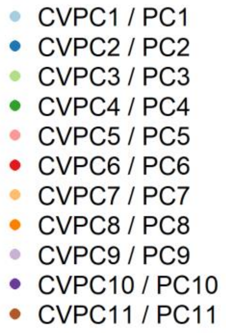


Integrative multivariate indices for biomarker variability (CVPCs, **A**) and levels (PCs, **B**) calculated every 3 months are plotted against time before death. Results from change point analyses applied to regression models between CVPCs and time before death, allowing slopes to vary across individuals, are indicated as dashed lines (**A**) and below the figure with 95% confidence intervals. All CVPCs except CVPC1 have very large confidence intervals, suggesting no clear change in their distribution before death.

**Supplementary Figure 11. Individual CVPC1 trajectories before death or censoring.**

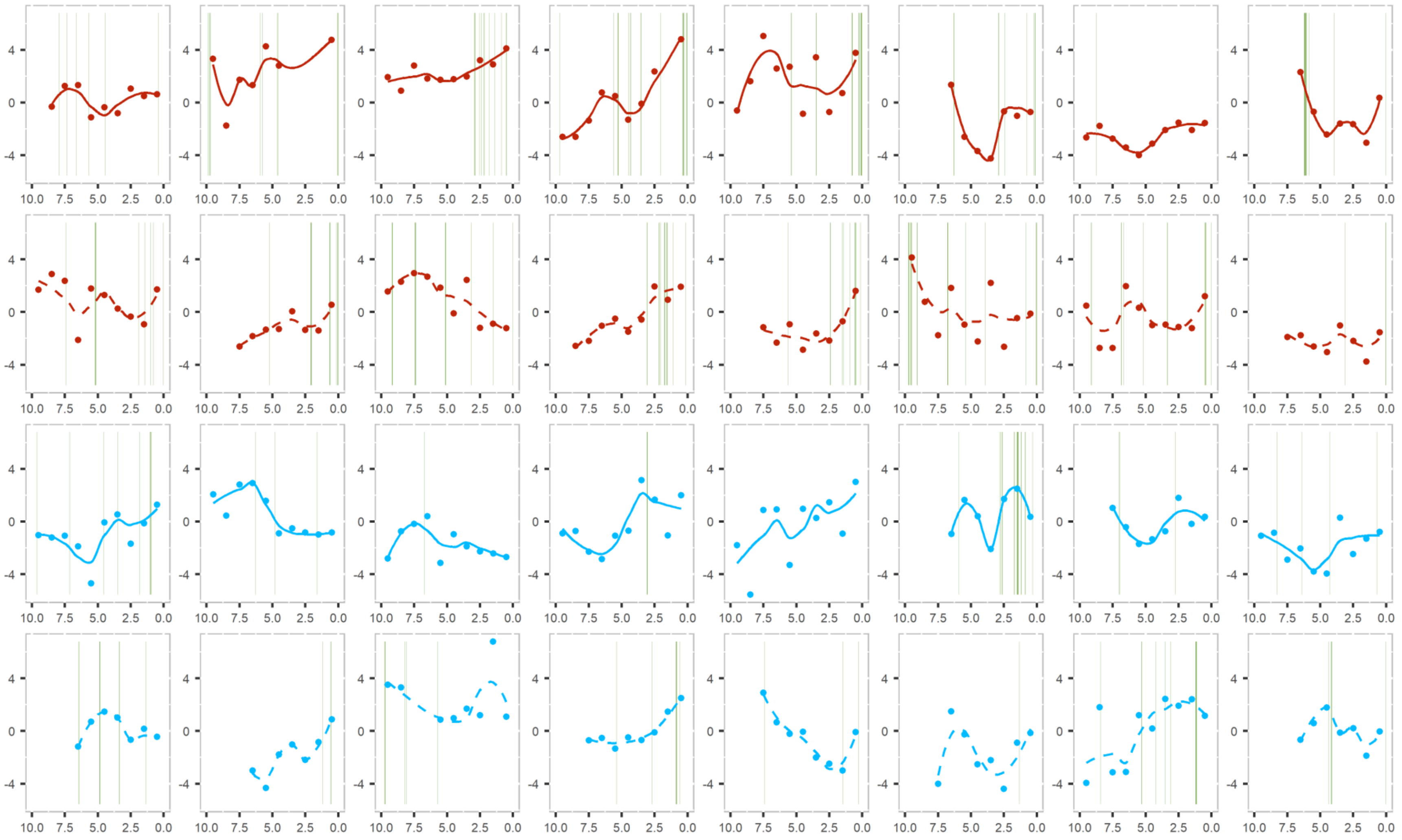

The color represents the status at the end of follow-up (red for patients who died and blue for patients who were alive) and line type represents the diabetes diagnosis (a solid line for non-diabetics and a dashed line for diabetic subjects). Vertical green lines represent hospitalizations. Individuals were randomly chosen from among those with ≥6 years of follow-up.

**Supplementary Figure 12. Mean and CV trends before death for the 4-month variable list.**

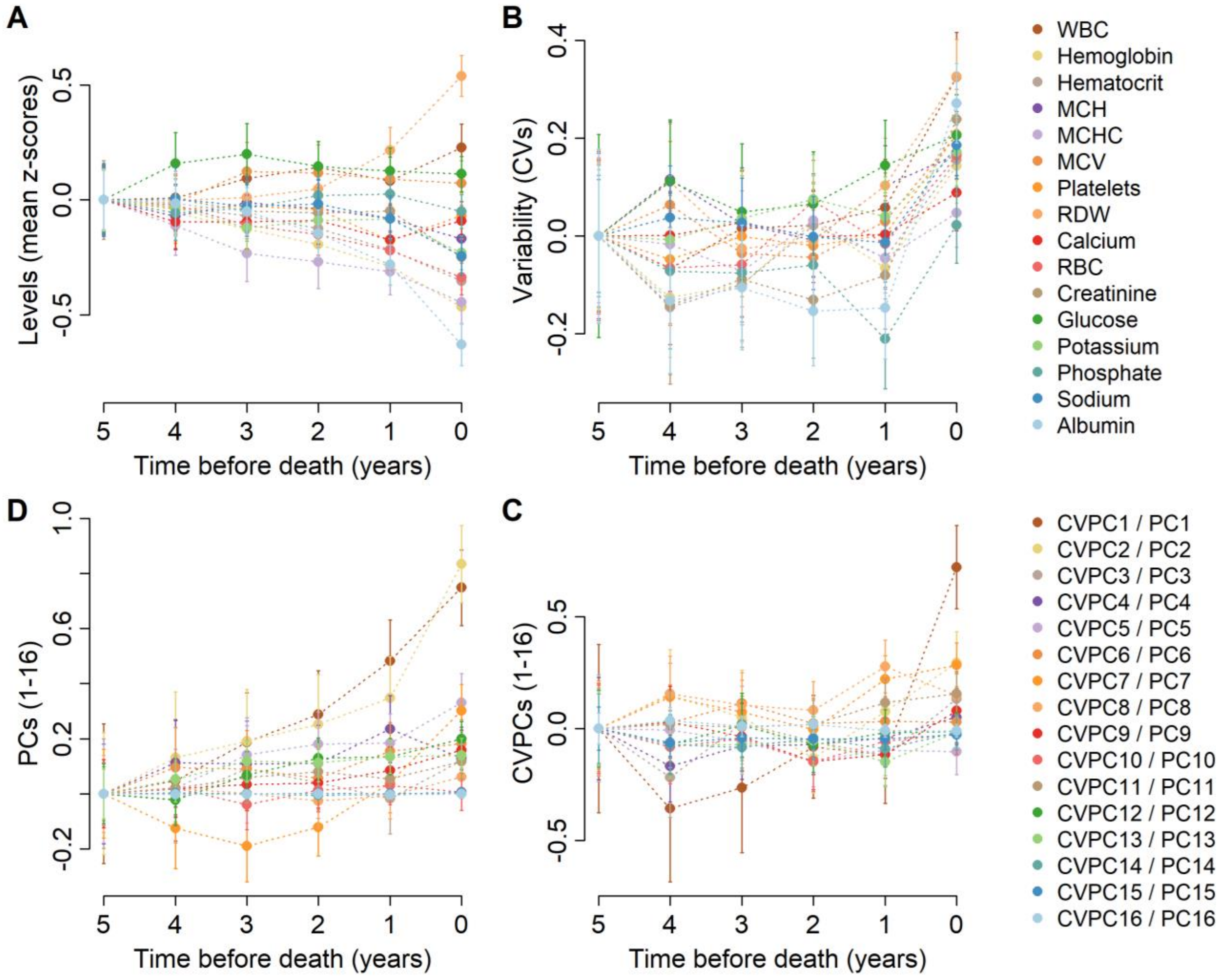


Biomarker levels (mean z-scores, **A**) and variability (CVs, **B**), as well as integrative multivariate indices for levels (PCs, **C**) and variability (CVPCs, **D**) were calculated every year and averages are plotted against time before death. For ease of comparison, means and CVs were centered at 5 years before death. Abbreviations: MCH, mean corpuscular hemoglobin; MCHC, mean corpuscular hemoglobin concentration; MCV, mean corpuscular volume; RBC, red blood cells; RDW, red cell distribution width; WBC, white blood cells.

### Supplementary Figure 13. Effect of the number of observations included in CV calculation.

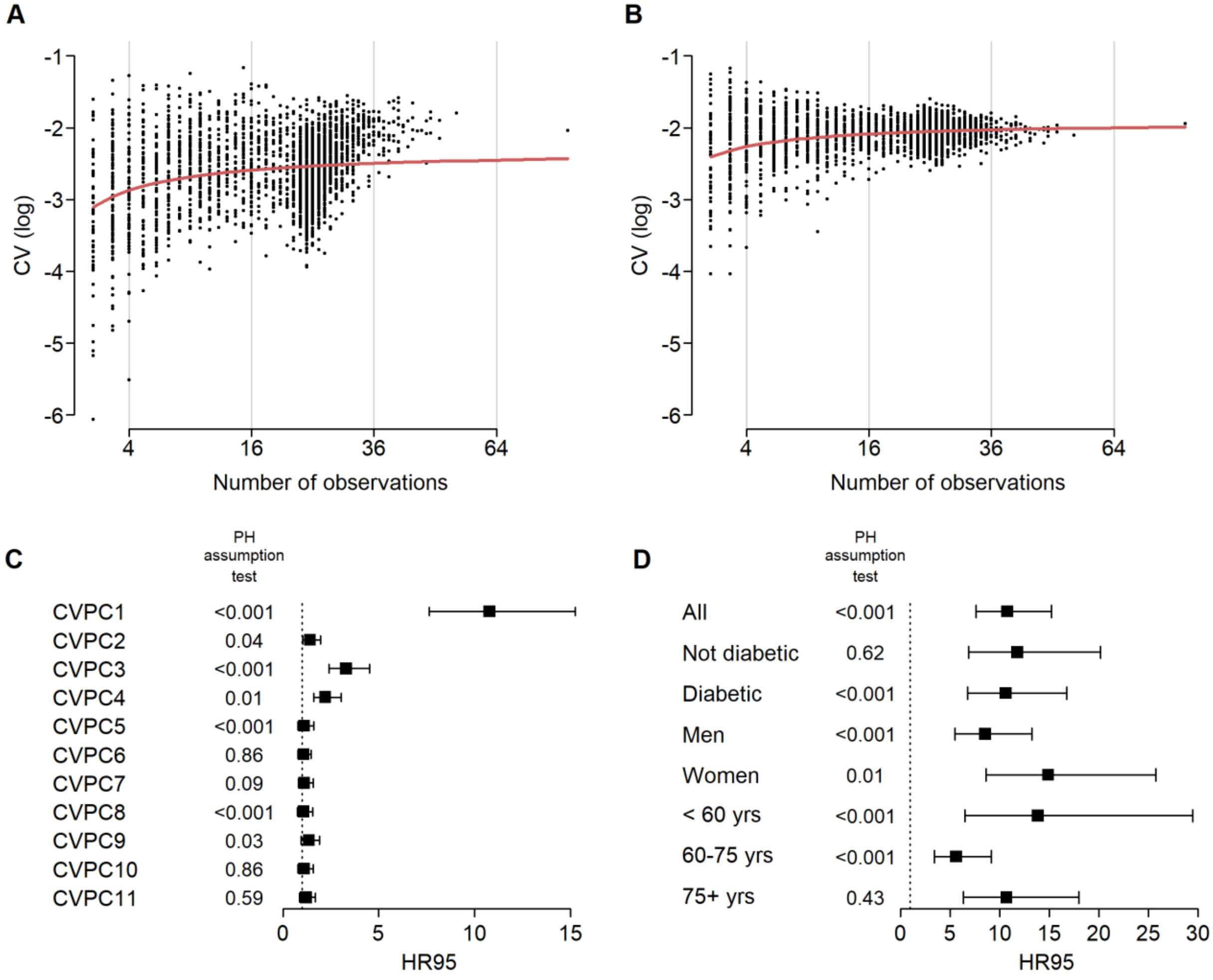


**A-B**, CV values tend to be smaller as fewer observations are included in its calculation, as observed in real data (**A**) and simulated data (**B**) for a given biomarker. All biomarkers show the same trend (data not shown). The red line represents the non-linear regression model used to correct for this bias, as described in the eMethods. **C-D**, HR95, the hazard ratio of being in the 97.5th percentile relative to the 2.5th percentile of the index, together with 95% confidence intervals are shown for CVPCs (**C**), as well as for CVPC1 calculated in different subsets of the study population (**D**), using only CVs calculated with at least 5 observations. All models control for age using a cubic spline (with 3 degrees of freedom when performed on a specific age group and 5 otherwise) and length of follow-up, clustering multiple observations per individual. Models also control for sex and diabetes diagnosis, except when population is stratified using this variable. These results are very similar to those obtained by correcting CVs for the number of observations included in its calculation (see Fig. 2 and Supplementary Fig. 2).
